# A Transcriptomic Atlas of the Human Brain Reveals Genetically Determined Aspects of Neuropsychiatric Health

**DOI:** 10.1101/2023.03.10.23287072

**Authors:** Xavier Bledsoe, Eric R. Gamazon

## Abstract

Imaging features associated with neuropsychiatric traits can provide valuable insights into underlying pathophysiology. Using data from the UK biobank, we perform tissue-specific TWAS on over 3,500 neuroimaging phenotypes to generate a publicly accessible resource detailing the neurophysiologic consequences of gene expression. As a comprehensive catalog of neuroendophenotypes, this resource represents a powerful neurologic gene prioritization schema that can improve our understanding of brain function, development, and disease. We show that our approach generates reproducible results in internal and external replication datasets. Notably, genetically determined expression alone is shown here to enable high-fidelity reconstruction of brain structure and organization. We demonstrate complementary benefits of cross-tissue and single-tissue analyses towards an integrated neurobiology and provide evidence that gene expression outside the central nervous system provides unique insights into brain health. As an application, we show that over 40% of genes previously associated with schizophrenia in the largest GWAS meta-analysis causally affect neuroimaging phenotypes noted to be altered in schizophrenic patients.

## Introduction

Several transcriptome-wide association studies (TWAS) have identified associations between genetically regulated gene expression (GReX) and neuropsychiatric traits. The contribution of GReX variation to disease pathophysiology remains an active area of research, with critical implications for our understanding of disease mechanisms^1–3^. Toward this end, we develop a publicly available resource, NeuroimaGene, for identifying the neurophysiological consequences of variation in GReX. We link 2 well-validated intermediate phenotypes: endogenous gene expression under genetic control and neurophysiology as captured by a broad array of neuroimaging derived phenotypes (NIDPs) from MR imaging^4–8^.

GReX characterizes the component of endogenous gene expression that is driven by germline variation in nucleotide sequence. This measure of genetic influence can be used to illuminate the biologic mechanisms of neuropsychiatric disease. GReX has been shown to be associated with a broad range of neuropsychiatric traits^9^. By quantifying the component of gene expression under genetic control, this measure is to be distinguished from environmental and trait-determined influences^8^. Since multiple SNPs often affect regulation of a gene, this transcription-targeted approach improves statistical power relative to single-SNP analyses by concentrating multiple SNP effects onto relatively fewer, biologically-informative (gene) targets^10^.

In this study, we leverage the UKB neuroimaging data to identify associations between variation in GReX and over 3,500 NIDPs. These quantitative descriptors of the structure, connectivity, and functional activation of the brain have been shown to be associated with neuropsychiatric health^11–15^. The UKB’s application of automated image processing pipelines to magnetic resonance imaging permits standardized measurements for an enormous number of cerebral data points^16–19^. These data capture structural information, such as the volume, thickness, and surface area of brain regions; organizational variation describing the white matter connections between these components; and functional information, such as networks of synaptic activity associated with specific tasks or resting states^15–18, 20^.

Through analysis of GReX in a population-based cohort, the resource captures the expression-mediated genetic architecture of structure and function in the healthy brain, facilitating the identification of specific genes and anatomic measurements involved in neurocognitive homeostasis^21^. In our application, we replicate and extend evidence characterizing the neurogenic pathophysiology of schizophrenia (SCZ).

## Results

### NeuroimaGene comprises a searchable atlas of associations between brain measures and the genetically regulated transcriptome

To identify associations between GReX and neuroimaging measures of the brain, we applied the JTI TWAS framework to summary statistics from the most recent release of the UKB neuroimaging GWAS (Figure 1a) (**Methods**)^5^. We used *in silico* gene expression models trained on transcriptome data in 19 tissues from the GTEx consortium. This analysis generated associations between 3,934 measures of the brain and 22,815 imputed gene expression traits in the context of 13 brain tissues and 6 neurologically relevant tissues from outside the central nervous system (CNS) (Figure 1d)^22, 23^. We removed non-heritable NIDPs and, recognizing the non-independence of NIDPs, we corrected for multiple testing through a study-wide Benjamini Hochberg false discovery rate threshold of 0.05. The majority of the significant associations represented white matter fibers identified via diffusion MRI and structural partitioning of the brain according to T1 imaging (Figure 1e). We provide these data as a searchable atlas detailing highly significant, quantitative associations between endogenous gene expression of >16,000 genes and >3,400 neuroimaging derived measures of the brain (Figure 1f) (See Data Accessibility).

**Figure 1:**
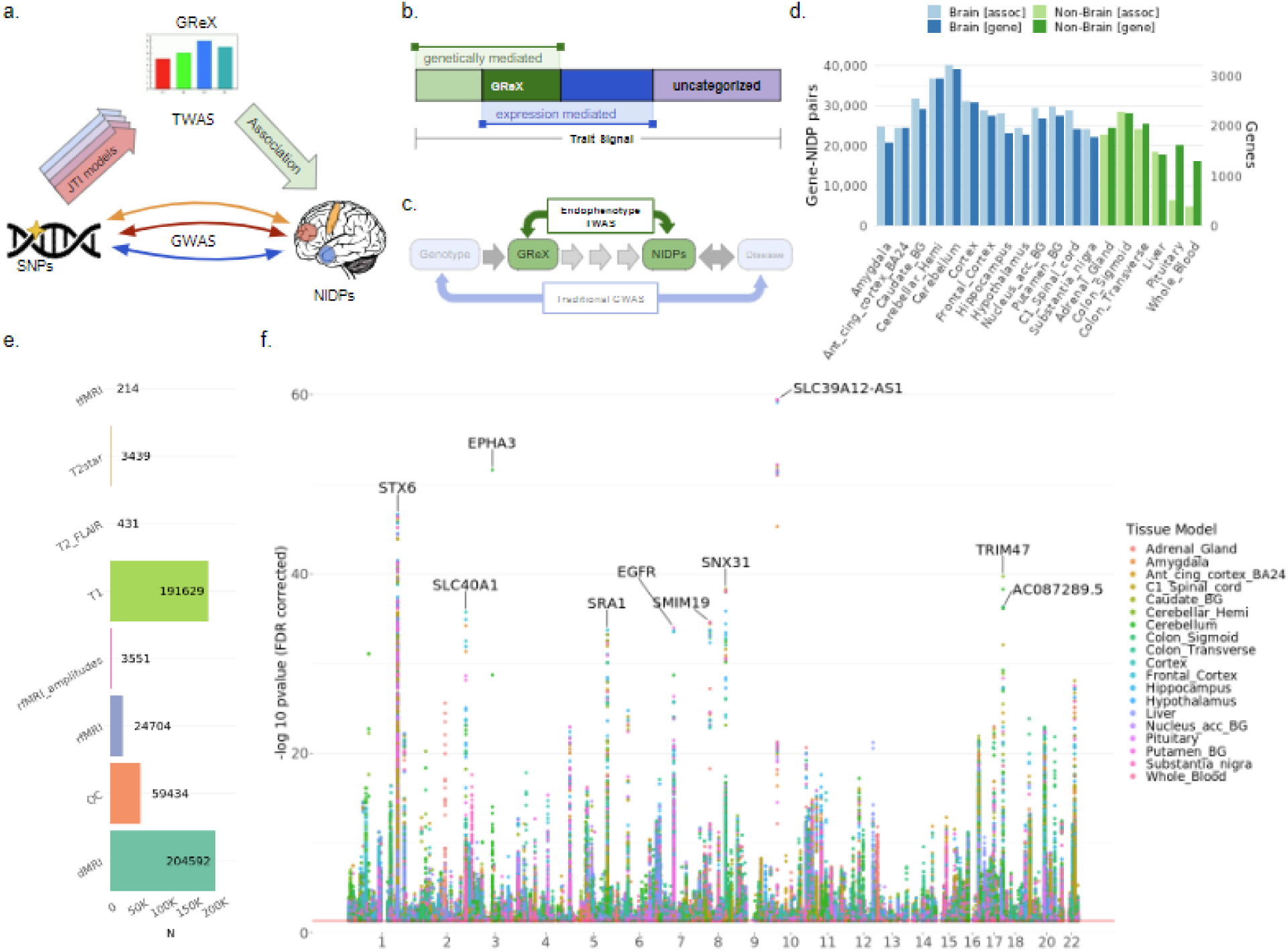
Resource Overview. **a**. Conceptual overview of the Neuroimaging TWAS framework leveraging the JTI-derived gene expression models. **b.** In relation to phenotypic risk, GReX can be used to quantify the fraction of SNV-based risk that is actualized via the intermediary of endogenous gene expression. **c**. The location of the endophenotype TWAS within the etiologic space strategically targets mechanistic elements mediating the associations between genetic variation and neuropsychiatric traits. **d.** The total number of GReX-NIDP associations identified in each tissue as well as the number of unique genes involved in those associations. **e.** Distribution of significant (FDR<0.05) GReX-NIDP associations across all tissue models grouped by the imaging modality used to capture each NIDP. **f**. Manhattan style plot of all GReX associations with NIDPs across the genome from the application of JTI models in 19 tissues. **GReX**: Genetically Regulated gene Expression, **SNV**: Single Nucleotide Variant, **TWAS**: Transcriptome-Wide Association Study, **GWAS**: Genome-Wide Association Study, **GTEx**: Genotype Tissue Expression consortium, **NIDP**: Neuroimaging Derived Phenotype, **eQTL**: Expression Quantitative Trait Locus, **JTI**: Joint Tissue Imputation.

### Clustering NIDPs according to GReX recapitulates the spatial patterning of the brain

We investigated the extent to which unsupervised phenotype clustering of all NIDPs based solely on GReX reflected known neurobiology. We performed hierarchical clustering of NIDPs according to the nominally significant GReX (Supplementary Figures 8-26). Notably, identical brain regions measured via different approaches clustered together and these clusters demonstrated close clustering with groups of physically proximal brain regions (Supplementary Tables 2-20). To better visualize the spatial patterning of the NIDPs, we applied UMAP to the cortical surface area and volume measurements^24^. Following optimization of parameters for global structure, annotation of points according to cortical area not only revealed clusters of NIDPs from the same regions, but also a global pattern that closely mimics the 2-dimensional, unfolded organization of the human cortex (Figure 2, Supplementary Figure 27). Subcortical regions clustered together near the brainstem and NIDPs describing the surface area and volume of different cortical regions self-aligned in both the rostral and longitudinal axes. While the three-dimensional cortex folds into itself at the longitudinal fissure, projecting the cortical surface into two dimensions should locate the deepest aspect of the fissure near the top of the image as is observed in the cingulate gyrus and corpus callosum. Classifying NIDPs exclusively by GReX thus provides sufficient data to identify a sample’s place of origin relative to other NIDPs, both underscoring the validity of GReX as a neurologically informative measure and highlighting the richness of information contained within the NeuroimaGene resource.

**Figure 2:**
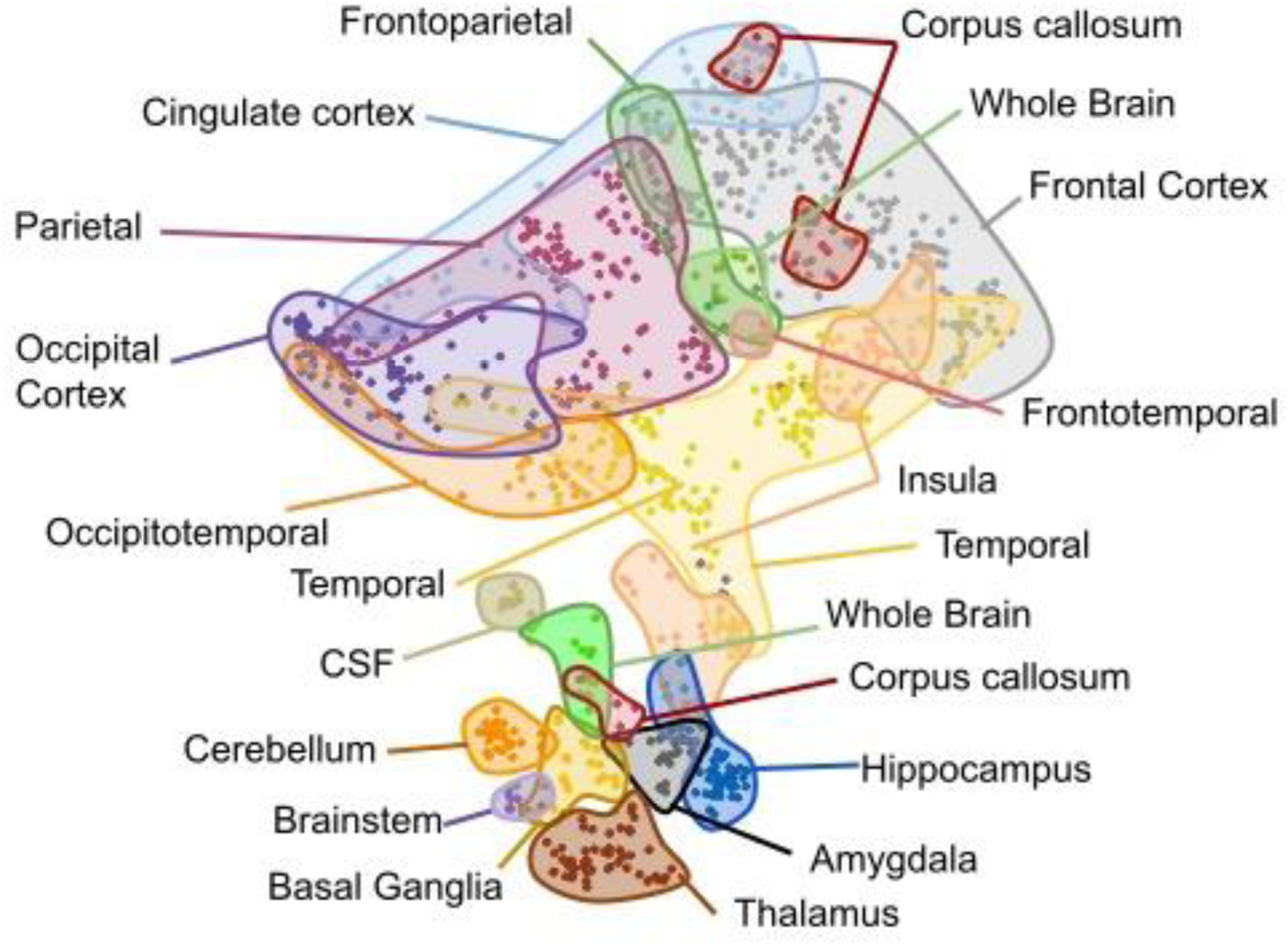
Effect of genetically determined transcriptome recapitulates brain structure and organization. UMAP projection of NIDPs corresponding to cortical surface area and volumes according to the Amygdala. The distribution of points in the cartesian plane is derived exclusively from Euclidean distance between the GReX effect size vectors for each NIDP. NIDPs are colored according to cortical region. Shaded areas delineated by hand.

### TWAS methodology for identifying neuroimaging GReX associations demonstrates high reproducibility in internal and external validation cohorts

The UKB neuroimaging GWAS release includes summary statistics of the full 33K subjects as well as two smaller GWA studies completed on populations subsets comprised of 22K and 11K non-overlapping sets of individuals. As methodological validation, we applied the TWAS pipeline to these smaller independent data sets (**Methods**). Using a nominal 0.05 p-value threshold cutoff, we identified 1,262,333 distinct GReX-NIDP associations, across all tissue-specific models from 19 tissues, that met the nominal threshold in the cohorts of 11K and 22K patients. These associations collectively represented 395,834 unique GReX-NIDP associations irrespective of tissue models. Notably, 75.8% of associations showed concordance in their direction of effect between studies (Supplementary Figure 30 a,b). The subset of study-wide significant findings (Benjamini Hochberg FDR < 0.05) demonstrated full concordance of direction of effect with an effect size Spearman correlation of 0.9812 (Figure 3a). Within each NIDP modality, the mean Spearman correlation across all NIDPs was always positive (Figure 3c). The number of nominally significant NIDPs across all tissue models in each modality category was largely skewed in favor of rfMRI, T1, and dMRI modalities (Figure 3b). Consistent with the heritability profile of NIDPs in these 3 modalities, the replication correlation was lowest for rfMRI but was reasonably high (> 0.5) for both T1 imaging and dMRI modalities (Figure 3d)^16^. In summary, our pipeline identified highly replicable associations between GReX and NIDPs across two independent, moderately powered data sets.

**Figure 3:**
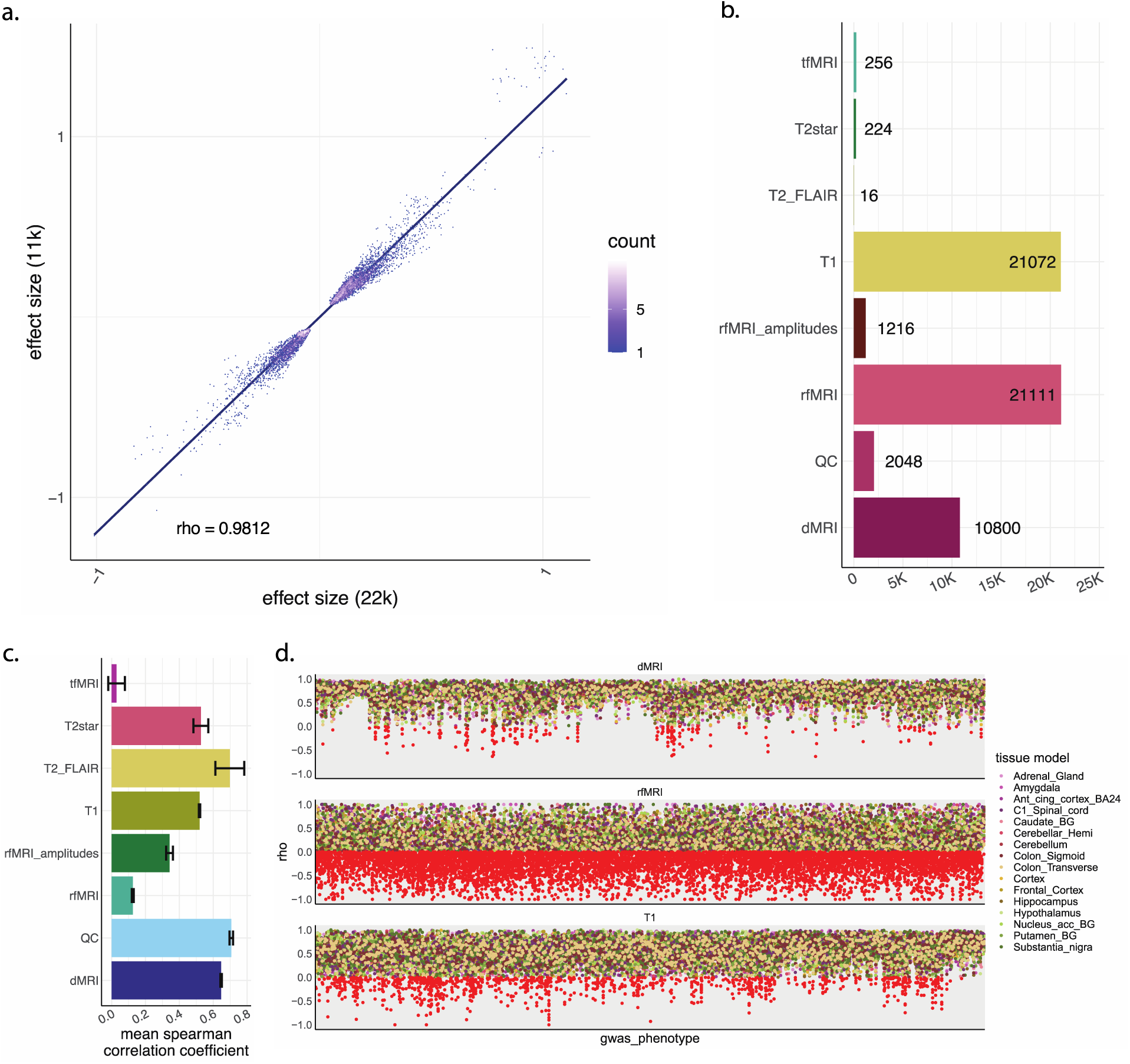
NeuroimaGene Methodology Validation. **a.** The correlation between effect sizes of tissue-concordant GReX-NIDP associations shared across the neuroimaging TWAS data from 11K subjects and the neuroimaging TWAS data from 22K subjects. Each point represents a study-wide significant finding (Benjamini Hochberg false discovery rate < 0.05). Data are plotted according to a pseudolog10 function with point color representing degree of overlap at each graphical coordinate. The trend line reflects linear regression. Spearman’s correlation = 0.9812 with p-value < 2.2e-16. **b.** Distribution of nominally significant NIDPs across all tissue models grouped by the imaging modality used to capture each NIDP. **c.** Mean Spearman correlation across all NIDP gene sets across data subsets grouped by the imaging modality used to capture the NIDP. Error bars represent the 95% confidence interval. **d.** Distribution of all NIDPs plotted according to the Spearman correlation coefficient between gene sets as calculated from TWAS in the 11K and 22K data subsets. Panels represent imaging modalities with only those representing dMRI, T1, and rfMRI shown. Point color represents the tissue model with all points showing anti-correlation filled red. (rho < 0) All data in subplots b-d filtered by a nominal p-value TWAS threshold of 0.05.

For replication of our findings in an external dataset, we applied the same TWAS pipeline used in NeuroimaGene to summary statistics from a cortical GWAS meta-analysis (N = 51,665) published by the Enhancing Neuroimaging Genetics through Meta-Analysis (ENIGMA) consortium^19^. The ENIGMA cortical GWAS identified SNP level associations with surface area and thickness measures of 34 different cortical regions. The consortium accessed raw MRI data from over 60 cohorts and performed cortical parcellation according to the Desikan atlas. We selected all FDR-significant associations from the Neuroimagene resource for the NIDPs that best corresponded to the phenotypes used in ENIGMA (see **Methods**). Matching these associations on tissue model, gene name, and NIDP, we replicated 47.6% of associations in the ENIGMA cohort using a nominal significance threshold (Supplemental Figures 33-34). This level of concordance was achieved in the presence of both diverse MRI acquisition protocols and differential in silico phenotyping of the NIDPs.

### Tissue specificity of neuroimaging GReX associations provides insight into the molecular pathways underlying neurobiology

We investigated the extent to which tissue context breadth provides meaningful information about GReX mechanisms. First, to characterize the distribution of GReX-NIDP associations across the 19 different tissue models, we annotated each gene according to the total number of tissue models in which the gene was significantly associated with any brain measure (Figure 4a). Here we relied on the FDR-significant subset of the data as is appropriate for discovery analyses. A plurality of genes demonstrated a statistically significant neurologic association in the context of only one single tissue model. Gene counts decreased as replication categories increased until reaching a nadir at 14. A similar pattern was observed when considering joint GReX-NIDP associations (Figure 4b). Expression of some genes was neurologically relevant in only a few tissues while for others, relevance for genetically determined expression was ubiquitous across tissue contexts. To explore the functional significance of this distribution, we conducted a comparative Gene Ontology enrichment analysis of tissue-specific genes against tissue-shared genes (Figure 4d) ^25^. GReX in both sets independently clustered into multiple ontology categories, indicating the non-random nature of the identified associations. The genes with tissue-shared GReX associations clustered into pathways that reflect housekeeping functions such as cell membrane integrity and mitochondrial function. Conversely, the genes with tissue-specific GReX associations included pathways reflecting cell projections such as those required for dendrite and axon formation and cell junctions. These categories illustrate the relevance of both general and highly cell-specific processes in the healthy development of the human brain.

**Figure 4:**
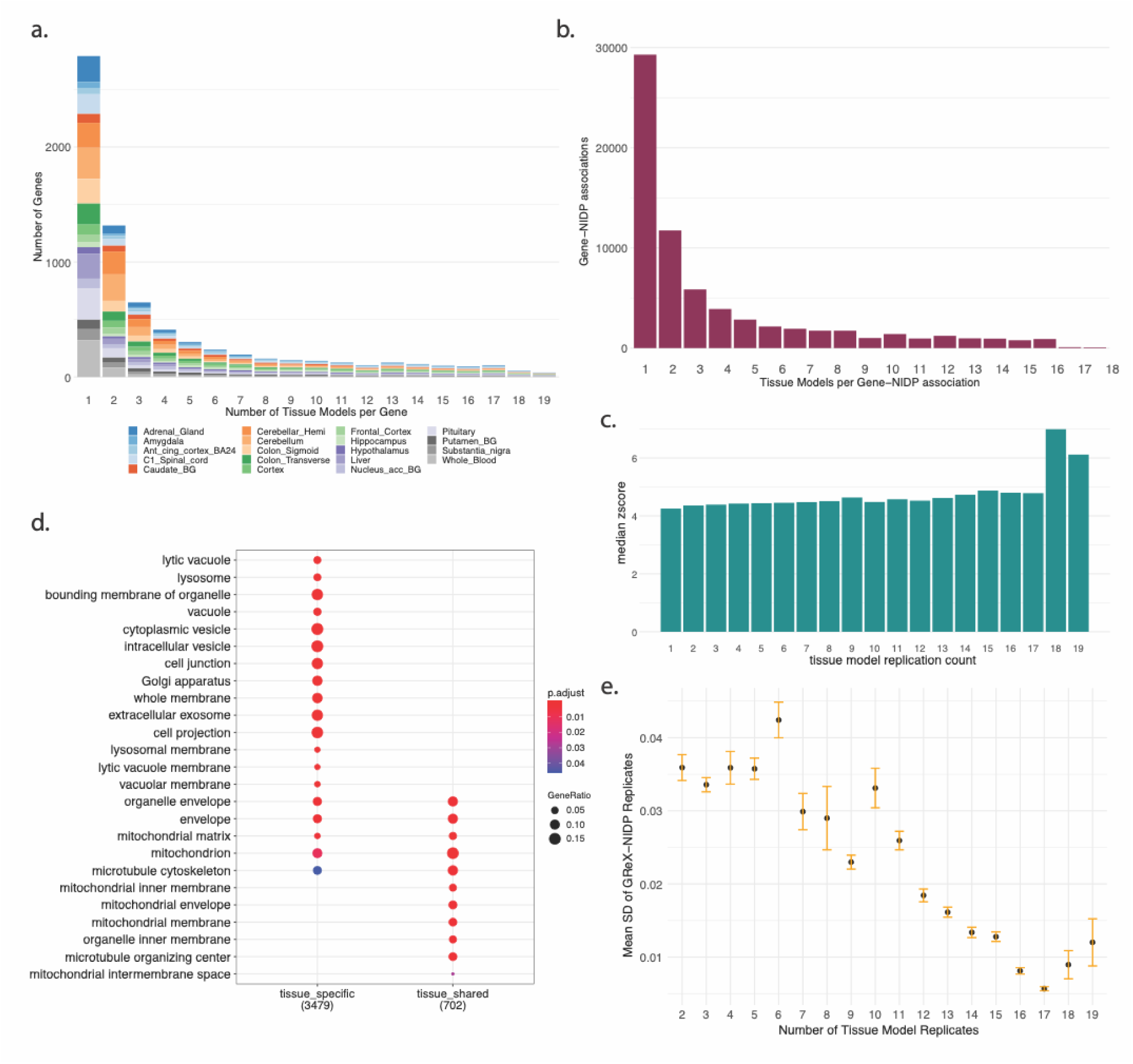
Tissue specificity of GReX is associated with magnitude of GReX effect and provides insights into underlying molecular pathways. **a.** Distribution of NIDP-associated GReX annotated on a scale according to the total number of tissue models (1 = unique to a single tissue model, 19 = shared across all tissue models). **b.** Distribution of GReX-NIDP associations annotated on a scale according to the total number of tissue models as in plot **3a**. **c.** Gene Ontology Enrichment Analysis of NIDP-associated GReX according to tissue model specificity. **d.** Median effect size of GReX-NIDP associations according to the total number of tissue models. **e.** Mean standard deviation of replicated GReX-NIDP associations across tissue models according to the number of tissue models. Error bars represent the 95% confidence interval.

We then assessed the relationship between the identified neurologic impact of gene expression and the number of tissues in which that expression is neuro-relevant through analysis of the mean effect size magnitudes across each category. We identified a positive relationship such that GReX-NIDP associations shared across all tested tissues showed relatively higher magnitude changes in neural architecture (Spearman rank correlation p = 7.73e-06, rho = 0.968) (Figure 4c). We calculated the standard deviation of effect size magnitude for each gene in each column and identified a negative relationship as tissue count replication increased (p-value < 5.33e-7, rho = -0.938) (Figure 4e). In summary, GReX-NIDP associations that were captured across multiple tissue models demonstrated greater effect size magnitude with less variance across tissue models.

These data in conjunction with the previous Gene Ontology suggested that tissue-shared GReX associations likely represent general biological processes that occur ubiquitously while those GReX-NIDP associations identified in a few tissues demonstrated more moderate, context-dependent effect sizes.

### GReX-NIDP associations identified in non-brain tissue models are highly relevant and provide information inaccessible from brain-only analyses

We assessed the significance of tissue model type (brain vs non-brain) with regard to the GReX-NIDP associations. As in the previous section, we annotated each gene according to the number of tissue models in which it was associated with a neurologic trait. Then we annotated the NIDP-associated GReX in each category according to the type of tissue model. The first tissue category, labeled as “non-brain”, consists of the adrenal gland, sigmoid colon, transverse colon, pituitary, liver, and whole blood. The rest of the brain phenotypes and the spinal cord are classified as “brain”.

Genes that showed significant associations exclusively in brain tissue models were classified as “unique-brain” while those that showed significant associations only in non-brain tissue models were classified as “unique-non-brain”. Those that demonstrated associations in both were labeled as “shared” (Figure 5a).

**Figure 5:**
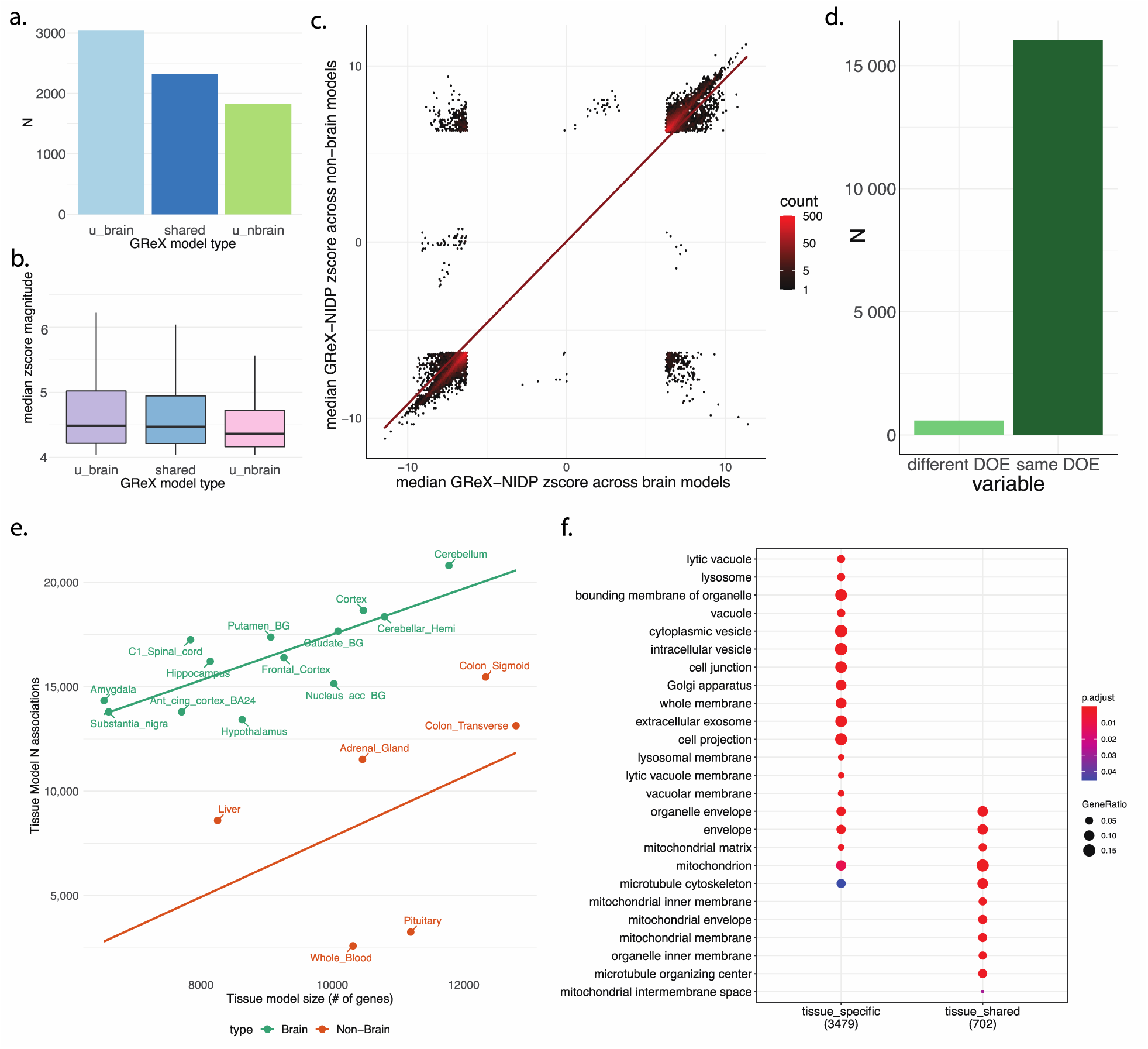
GReX in both brain and non-brain tissues provides complementary information reflecting the integrated nature of neurobiology. **a.** Bar plot showing the counts of all 7,193 GReX classified according to their presence in brain tissue training models, non-brain models, or both. **b.** Box plot showing the median normalized effect size across all significantly associated NIDPs and the interquartile range for all 7,193 FDR-significant GReX grouped in the same manner as in panel a. **c.** Density plot showing the distribution of median normalized GReX-NIDP effect sizes as measured in brain tissue models vs the same measure as captured in non-brain tissues. Only GReX-NIDP associations shared across both types of tissue models are represented here (N = 1,262,333, FDR corrected p-value < 0.05) trend line represents a linear regression. **d.** Relative counts of tissue-concordant GReX-NIDP associations that share the same direction of effect across tissue types vs those that show differing directions of effect. **e.** The number of GReX associations per tissue model plotted against the number of genes in the tissue model, grouped by model type (brain vs non-brain). Trend lines represent linear regression. **f.** Gene Ontology Enrichment Analysis of NIDP-associated GReX according to tissue model type.

As expected, the most informative GReX-NIDP associations were identified by models derived from brain tissue. Still, nearly 50% of GReX associations detected in brain tissues were also detected in non-brain tissues, underscoring the potential for non-brain tissues to serve as proxy models for the hard-to-access CNS. Accordingly, for each “shared” GReX-NIDP association, we assessed the similarity in estimated effect size across brain and non-brain groups by comparing the median effect sizes of the 2 tissue groups (Figure 5c). GReX-NIDP associations captured across both brain and non-brain models demonstrated high consistency in cross tissue-type effect size directions (Figure 5 c,d).

We noted with interest that there were over 1700 genes for which GReX exclusively outside of the brain was significantly associated with neurophysiological traits (Figure 5a). This seemed to suggest that transcriptomic variation outside the CNS was highly relevant to brain physiology. The median effect size magnitude of GReX associations in non-brain tissues was significantly lower than that of brain tissues (p-value < 2.2e-16); however, the difference was small with the median effect size falling within the interquartile range for the brain data (Figure 5b).

Each JTI tissue model involves a variable number of genes. The ratio of total neurophysiological associations derived from each tissue model to the number of genes in each tissue model thus reflects a measure of marginal information gained per additional gene. We compared these ratios among the tissues (Figure 5e), demonstrating that the amount of information gained by increasing the size of non-brain tissue models was statistically no different than the information gained by increasing the size of brain tissue models (p-value = 0.949). While brain tissue retained an absolute advantage over non-brain tissues, the marginal gain in neurophysiological information from each new gene was the same across brain and non-brain models.

To characterize the functional ramifications of this geographic partitioning, we conducted comparative Gene Ontology analysis, contrasting the “unique brain” NIDP-associated GReX with the “unique non-brain” NIDP-associated GReX (Figure 5f)^25^. For genes exclusively associated with NIDPs via brain expression models, there was again heavy enrichment in cell projection pathways as are involved in axonogenesis and dendrite formation. In non-brain tissues, there was an enrichment in pathways reflecting cellular metabolism and cell signaling.

### NeuroimaGene application to schizophrenia recapitulates and extends upon previously documented observations

We modeled the utility of the GReX-NIDP association resource through an application to SCZ. Remarkable in both its high SNP-based heritability and polygenicity, the most recent GWAS meta-analysis identified 263 trait associated loci with mapped variants supporting a narrow sense heritability of 60-80%^26^. Recognizing the joint influence of germline variation and neurologic predisposition, we used NeuroimaGene to identify relationships between GReX of SCZ associated genes and SCZ associated NIDPs. We gathered 33 SCZ-associated cortical NIDPs and 12 subcortical volumes identified by the ENIGMA consortium that were present in the UKB Neuroimaging Study (**Methods**)^27, 28^. We then identified 63 SCZ trait-associated genes from a TWAS analysis conducted on the most recent and largest SCZ GWAS meta-analysis to date (**Methods**)^26^. Of these genes, 50 were present in the tested GReX of the UKB TWAS analysis. Accordingly, we queried the NeuroimaGene resource for associations between GReX of the SCZ associated genes and SCZ-associated NIDPs (Figure 6a).

**Figure 6:**
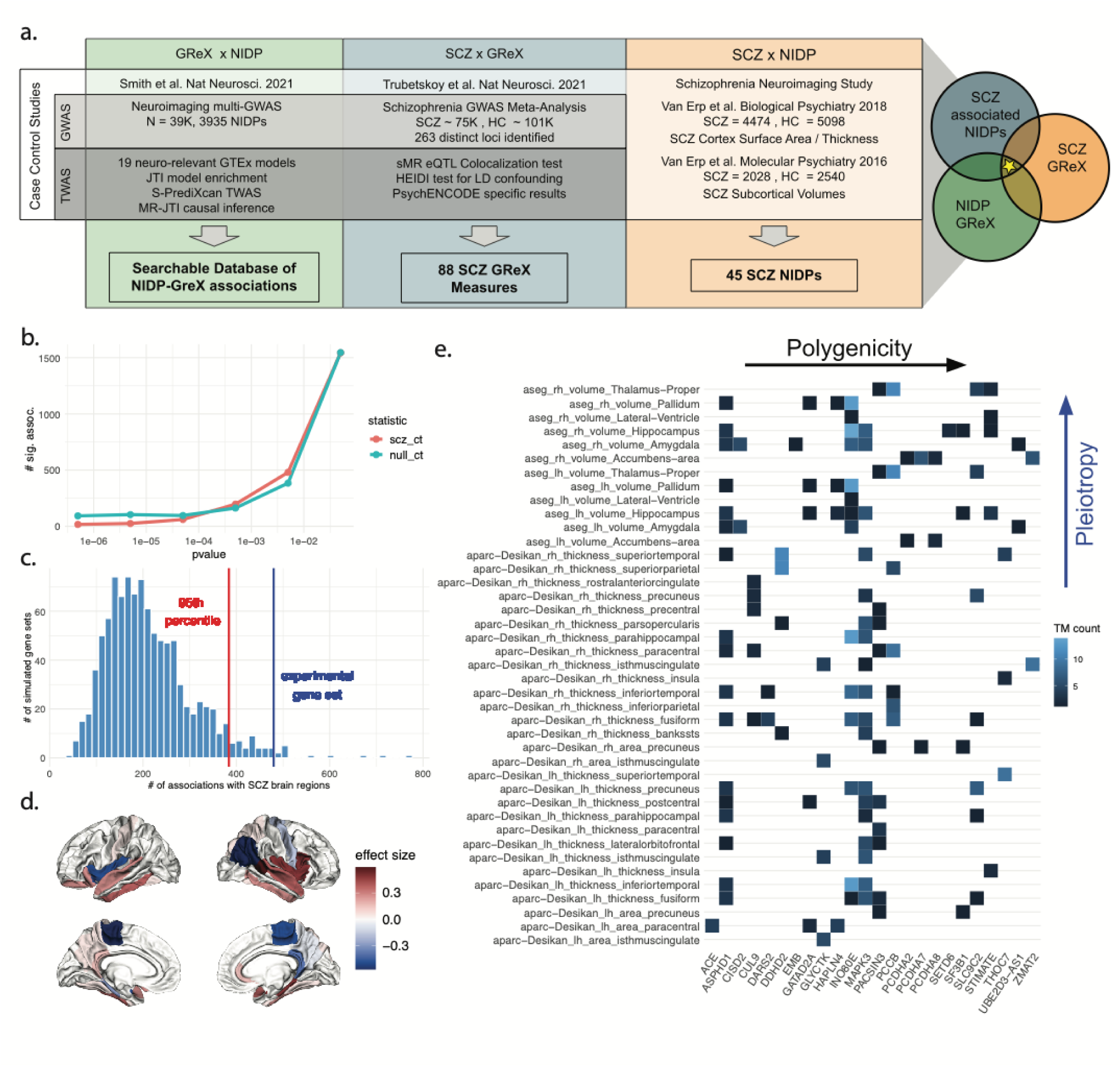
NeuroimaGene application to SCZ informs clinical neuroimaging findings. **a.** Schematic overview of the application of the pipeline to identified tri-fold associations between GReX, SCZ and NIDPs. **b.** Comparison of total associations between SCZ GReX and SCZ NIDPs (scz_ct) and total associations between random GReX and SCZ NIDPs (null_ct) at different p-value thresholds. **c.** Quantification of SCZ GReX associations with SCZ NIDPs compared to a null distribution at p-value <= 0.005. The experimental gene set represents the set of GReX measures associated with SCZ according to Trubetskoy et al. The histogram represents the distribution of associations between 1000 random equally sized gene sets and SCZ NIDPs. The line in red represents the limit of association counts exceeded b only 0.05 of the random gene sets. The blue line represents the number of total associations between SCZ-associated NIDPs. **d.** Visualization of the surface area and volumetric consequences of GReX for schizophrenia-associated genes as detected in the Desikan Atlas using fsbrain29. The effect size represents the mean GReX-NIDP effect size corrected for LD contamination via Mendelian Randomization (MR-JTI). The intensity of the effect size represents the magnitude of the effect size and the color represents the direction of effect of increased GReX on the size of the region in question (blue = negative, red = positive). **e.** Distribution of MR-JTI significant associations between GReX of SCZ GReX and SCZ-associated NIDPs with significance values <= 0.005. Polygenicity of individual NIDPs within the set of SCZ GReX can be observed across the x-axis. The pleiotropic effects of SCZ GReX on SCZ NIDP’s can be observed along the y-axis

### SCZ NIDPs are more strongly associated with GReX of SCZ associated genes relative to the full imputed transcriptome

We expected that GReX of the trait-associated genes identified by the SCZ TWAS would be more associated with the set of SCZ NIDPs than GReX of random genes, owing to the co-association of both variables (GReX and NIDPs) with SCZ status. Identifying an appropriate significance metric is complicated as GReX is highly pleiotropic with regard to brain measures and the NIDPs are largely polygenic. This high level of association between GReX and NIDPs represents the background against which the known SCZ associations must distinguish themselves. Having previously demonstrated that there was significant information in the resource using a nominal threshold (Figure 2), we set 0.05 as an upper limit and then identified the p-value threshold that would maximally discriminate between SCZ NIDP-associated GReX and the rest of the genome by testing a range of p-values, as is often done in polygenic risk score development.

We explicitly tested the null hypothesis that SCZ genes do not have more statistically significant associations with SCZ NIDPs (across all tissue models) than random, equally sized sets of genes. We conducted permutation testing and repeated the analysis across a range of p-values (**Methods**). We compared the value of the observed test statistic against the 95th percentile of the null distribution (Figure 6b-c, Supplementary Figure 35). We then calculated all significant SCZ-GReX associations with SCZ clinical NIDPs using 0.005 as the optimal p-value.

Following causal inference analysis via MR-JTI (a Mendelian randomization framework)^5^, we identified 467 causal associations between GReX of SCZ associated genes and clinical SCZ-associated NIDPs (Figure 6d-e, Supplementary Figure 36). These associations highlighted 42 NIDPs reflecting cortical and subcortical morphology and involved 25 genes. These two classes of intermediate variables were previously associated with SCZ (GReX and NIDPs), and we demonstrated in the UKB data that they exist in a causal relationship with one another whereby GReX drives the measured variation in morphology.

We observed notable degrees of polygenicity and pleiotropy with regards to the NIDPs and GReX, respectively. The GReX of all 25 SCZ associated genes showed association with an average of 5.2 SCZ-associated NIDPs. Each NIDP was associated with an average of 3.0 NIDP-associated genes (Figure 6e).

## Discussion

In NeuroimaGene, we provide a publicly available resource for examining neurological correlates of genetic variation. The number of studies identifying associations between genetic variation and neuropsychiatric traits is constantly increasing and so too does the importance of enriching these observational studies with data on a wide array of intermediate phenotypes. By characterizing the impact of genetically regulated gene expression on diverse measures of brain structure and function, we provide a tool that is broadly applicable to questions of development, neural maintenance, neurodegeneration, cognition, and psychiatric health.

The resource leverages joint-tissue imputation enriched models which have been demonstrated to outperform traditional single-tissue models. By analyzing multiple tissues rather than a single cross-tissue analysis, we retain information on the geographic partitioning of predictive and causal GReX variation. This is enhanced by the inclusion of non-brain tissues. The breadth of NIDPs characterized by the UKB and subsequently described here represents a major advantage of this resource as well. There is substantial variation in how different cortical, subcortical, and DTI atlases parcellate the brain. Because NeuroimaGene includes a wide breadth of such atlases, conversion between atlases is rarely necessary when searching for GReX associated with an NIDP of interest.

The utility of GReX as an informative endophenotype for neurologic features is strongly underscored by the clustering analyses wherein we recapitulate the 2-dimensional structure of the human cortex and subcortex, using effect of the genetically determined transcriptome alone. The highly significant replication of associations across the two independent population subsets and a third external data source further demonstrates the strength of our approach. Additionally, stratifying the neuro-relevant genes according to tissue specificity followed by Gene Ontology analysis demonstrated that tissue specificity approximates differentiation. While associations shared across tissues, as are prioritized by cross-tissue analyses, capture a high-confidence set of associations with larger and more uniform effect sizes, they miss some associations captured in single-tissue analyses that are reflective of greater differentiation/specificity.

Clinically, the link between corporeal health and neuropsychiatric health is well-established with conditions such as hepatic encephalopathy and pheochromocytoma-induced panic disorder representing high visibility instances. Using 6 non-brain tissues with a priori evidence for neurologic relevance, we present data suggesting that transcriptomic variation in tissues outside the brain has important effects on neurologic homeostasis that are large enough to be captured by neuroimaging methods.

By prioritizing SCZ-associated GReX, these data support a genetic basis for cortical morphology changes observed in SCZ. Furthermore, we provide specific gene candidates for which expression variation may be driving these cortical changes.

This work is limited by several factors. First, regulatory genomic architecture and linkage disequilibrium scores translate poorly across populations of different ancestral backgrounds. In relying on genetic data derived from the fraction of genetic diversity contained in European ancestry, these findings are severely limited in their wide-scale application to human populations. In future studies we hope to leverage a broader range of data as reference transcriptome resources become available for less well-characterized populations. Second, the use of MR controls for horizontal pleiotropy at the level of SNPs; however, we do not control for confounding that is mediated through genetic co-expression^30^. Though mitigated by using cis-eQTL models, this limits the rigor of causal inference and necessitates the use of functional studies to validate findings.

SNP variation can affect a phenotype through many different mechanisms. By measuring the impact of SNPs only on gene expression, our approach does not capture the proportion of narrow-sense heritability mediated through other mechanisms^9^. This loss in breadth is redeemed by the increase in mechanistic specificity. Because our source data comes from healthy patients, neuroanatomical changes exclusive to disease-causing variants will be poorly covered^7^. Because GReX is a continuous exposure, we anticipate that some neurological predisposition will be present in the brains of individuals with non-realized genetic risk.

## Methods

### Data acquisition

The UK biobank (UKB) is a large-scale prospective cohort study, which has collected genotypic and phenotypic data from 500,000 residents of the United Kingdom. The standardization of data collection methods across patients and the large sample size of this study provides enormous advantages for epidemiological analysis. In 2014 the UKB began collecting neuroimaging data on patients in the form of functional, diffusion, and structural MRI^7^. The study developed an automated image processing pipeline to standardize and quantify their findings, generating 3,935 ‘image derived phenotypes’^20^. In 2016, Elliot et al published a GWAS for each of the 3,144 image derived phenotypes using genetic information from 8,428 patients available in the UKB database at the time^17^. In 2021, an additional release of neuroimaging GWAS data was made available detailing associations for 3,935 NIDPs as measured in 39,691 brain imaged samples derived from approximately 33,000 patients^16^. The summary statistics results for these GWAS are publicly available at their website (https://open.win.ox.ac.uk/ukbiobank/big40/).

### Statistical analysis

All analyses are conducted in R version 4.0.5, Python Python 3.8.6, and Linux.

### Model building and JTI framework

We leveraged pre-built gene expression models trained on RNA data from post-mortem samples in order to predict gene expression from SNP profiles. The construction of these models is described in Zhou et al 2020^5^. The GTEx consortium maintains a repository of RNA expression data and genetic data collected from 54 non-diseased tissue sites across approximately 1000 individuals^31^. As described in Zhou et al, each model quantifies the association between SNPs and gene expression levels across the genome for each tissue. The genetic regulation of gene expression is highly shared across tissues and JTI exploits this shared regulatory architecture to substantially improve expression prediction. These models, trained on the GTEx data and enriched via JTI, are publicly available at Zenodo (see Data Availability).

### TWAS of neuroimaging phenotypes

We applied the JTI TWAS models derived from 19 GTEx (version 8) tissues to UKB GWAS summary statistics for all 3,935 NIDPs published by Smith et al. to identify GReX associations with NIDPs^4^. We used S-PrediXcan, which generates the TWAS summary statistics. The methodology incorporates variance and covariance data of SNPs according to a linkage disequilibrium reference panel.

### Multiple testing correction in discovery cohort

Our study analyzed 22,815 genes and 19 GTEx-derived JTI tissue expression models across 3,935 NIDPs. We adjusted for the multiple hypothesis testing using the Benjamini-Hochberg false discovery rate threshold of 0.05. For all analyses involving significance testing, we considered only the subset of NIDPs with significant SNP-mediated heritability (3,546)^16^.

### Gene set analysis

We conducted GO term enrichment analyses using the ClusterProfiler R tool^25^. For all GO analyses, we set the size range for gene sets as 15-500 and used a Benjamini Hochberg p-value cutoff of 0.05 for filtering of pathway associations. The Cellular Components library of Gene Ontologies was used.

### Correlation analysis of TWAS results

We estimated the correlations in TWAS associations among the NIDPs. We first stratified the TWAS associations according to the tissue model. Each tissue model was represented by a matrix detailing the normalized effect size (z-score) of each gene on each NIDP. Each matrix consists of ∼22,000 rows (genes), 3,934 columns (phenotypes), and each data point is the normalized effect size (z-score) of GReX on the NIDP. Instances where specific genes were not associated with certain phenotypes were coded as NA. Concerning data completeness, we used a 99% completion threshold for the inclusion of phenotypes in the tissue specific NIDP clustering analyses. No phenotypes failed thresholding. Using the matrix for each tissue model, we calculated the pairwise Spearman correlation for all NIDP pairs according to the TWAS z-scores.

After identifying the Spearman correlation distance between all NIDPs according to the z-scores from the GReX effect, we reproduced the data in the form of a heatmap with axes ordered by ward’s agglomerative hierarchical clustering method (Supplementary Figures 8-26). The NIDPs involved in this ordering can be categorized according to a variety of criteria, including imaging modality, atlas used, lobe of the brain (for cortical measures), direction of fibers (for white matter tractography), left or right hemisphere, etc. To classify the NIDPs according to these categories, we annotated each NIDP according to the multi-dimensional phenotypic descriptors provided by the UKB (Supplementary table 1). These annotations allow for visual annotation of each of the elements in the clustered correlations. Using the R package *complexheatmap*, we created annotated, clustered heatmaps for all the phenotypes according to z-scores for each tissue model.^32^

### UMAP dimension reduction of T1 and dMRI phenotypes

We applied the Uniform Manifold Approximation and Projection for Dimension Reduction (UMAP) package in R to improve the visual interpretability of clustering analyses for 2 subsets of GReX-NIDP associations^24^. Replicated across all 19 tissue models, we conducted UMAP analysis on the GReX measures nomically associated with (1) T1-weighted cortical and subcortical surface area/volume measures (Supplementary Figure 27) and (2) dMRI tractography measures (Supplementary Figure 28). We optimized the parameter values for n_neighbors (number of approximate nearest neighbors used to construct the initial high-dimensional graph), and min_dist (minimum distance between points in low-dimensional space) in order to prioritize global structure over local structure. This was performed through iterative analyses of each data subset across multiple combinations of the two parameter values and visual evaluation of cluster projection. Final parameter sets were selected based on those which generated the most interpretable separation and clustering between NIDPs according to annotations of side, region, and type of measurement. For the T1 dataset, we selected n_neighbors = 150 and min_dist = 0.09. For the dMRI dataset, these values were n_neighbors = 60 and min_dist = 0.1. We set initial seed values for the R-session as well for the UMAP internal functionality to improve the reproducibility of the analysis (see Code Availability).

### PCA dimensional reduction of T1 phenotypes

We conducted Principal Component Analysis of the T1 structural NIDPs in R according to the normalized effect size of each GReX-NIDP association. Pooling data from all 19 tested tissue models, we displayed the first two principal components of the data (Supplementary Figure 29). We annotated the NIDPs by color with variation representing the type of cortical measure (surface area, thickness, or volume.)

### Internal replication analysis for neuroimaging TWAS

The UKB generated the Neuroimaging GWAS in 33K patients. In addition to this large data set, the UKB released 2 mirror versions following the same analytic pipeline but using 2 independent subsets of the populations with sample sizes of 11,000 and 22,000. To test the replication of the Neuroimaging TWAS methodology, we replicated our initial TWAS in these two smaller data sets for independent comparison. We applied the 19 JTI TWAS models to the 11K dataset and the 22K dataset independently. In order to compare these two TWAS datasets, we took a conservative approach and selected all associations from both studies that meet a nominal p-value of 0.05. From these data, we selected the GReX-NIDP associations that are present in both the 11K and 22K cohorts (matched on JTI TWAS models). These data represent TWAS associations between GReX and NIDPs derived from independent cohorts curated by the UKB. We present all these associations in Figure 3a with the effect size as derived from the 11K cohort on the y axis and from the 22K cohort on the x axis. We calculated the (non-parametric) Spearman correlation as effect sizes were not normally distributed.

We identified the direction of effect (-1, 1) for each association in each cohort and identified the distribution of shared associations with concordant direction of effect vs discordant direction of effect (Figure 3b). Using the MRI modalities described in the Neuroimaging GWAS supplement, we assessed the distribution of all shared associations grouped by modality (Figure 3c).

We calculated the mean Spearman correlation between the 11K cohort and the 22K cohort for each NIDP. These data were presented by modality in Supplementary Figure 31. We then took the mean of all NIDPs in each modality category to identify the mean Spearman correlation value for each NIDP per modality (Figure 3d). We replicated this analysis, dividing the data by both modality and JTI model (Supplementary Figure 32).

### External replication analysis of neuroimaging TWAS in ENIGMA

The Enhancing Neuroimaging Genetics through Meta Analysis (ENIGMA) consortium has published a GWAS meta-analysis of cortical measures derived from 51,665 individuals.^19^ The phenotypic measures include the surface area and thickness of the whole cortex as well as 34 cortical regions as defined by the Desikan cortical atlas.^33^ We accessed the version of the data describing regional surface area and thickness that had been adjusted for the respective global measures and undergone genomic control adjustment to account for population stratification across the many different participating sites in ENIGMA. We then applied S-PrediXcan to the subset of ENIGMA NIDPs that demonstrated significant SNP based heritability from the initial GWAS. We used the same 19 tissue-specific, JTI gene expression models described for the UKB neuroimaging TWAS. We filtered these GReX-NIDP-tissue associations according to a minimum r^2^ of 0.1 and a nominal p value threshold of 0.05.

The ENIGMA consortium consolidated the left and right hemispheric regions into single phenotypes prior to performing the GWAS analysis. To best match the phenotypes between the two studies, we first identified all the TWAS associations from the UKB neuroimaging study that matched the heritable cortical area and thickness measures described in ENIGMA irrespective of hemisphere. Then for NIDPs that were captured bilaterally in the UKB but not in the ENIGMA study, we selected genes whose GReX was significantly associated with the NIDP of interest in both the right and left hemisphere. This resulted in a set of hemisphere agnostic, tissue-specific GReX-NIDP associations for the Desikan cortical regions derived from the UKB data. We then calculated the proportion and explored the distribution of tissue specific GReX-NIDP associations from the UKB discovery dataset that achieved nominal significance in the ENIGMA replication dataset.

### Schizophrenia-associated GReX curation

We identified GReX associated with SCZ using the most recent major SCZ GWAS meta analysis from Trubetskoy et al^26^. This extended GWAS consists of the core Psychiatric Genomics Consortium dataset, an African American dataset, a Latino dataset, and the deCODE SCZ cohorts. Sample details are provided in the Supplementary information for the initial publication. Trubetskoy et al performed secondary analyses on the findings from the extended GWAS using patients from the core PGC cohort owing to accessibility of individual level data. Of relevance to this work, they performed Summary Mendelian Randomization on the extended GWAS findings to highlight 101 unique genes associated with SCZ through genetically regulated expression. For TWAS models, they leveraged those derived from PsychENCODE, eQTLGen, and fetal brain data. For the sake of uniformity in data analysis, we selected the subset of 88 genes identified via SMR using the PsychENCODE data. These data included GReX associations from eQTL models trained on brain and whole blood. Only the GReX from the whole blood analysis that were also replicated in brain models were reported by Trubetskoy et al. for the sake of prioritizing neurological relevance. Of these 88 GReX, 63 were able to be mapped to unique Ensembl IDs, 50 of which were present in the NeuroimaGene resource.

### Schizophrenia NIDP curation

SCZ NIDPs were derived from two studies published by the ENIGMA consortium, one highlighting cortical regions and the other subcortical regions^27, 28^.

In assessing cortical changes associated with SCZ, van Erp et al analyzed patient data from 39 different centers across the globe. These data represent 4474 individuals with SCZ and 5098 healthy controls, all of whom were assessed using a standardized approach. Cohort details and analysis methods were described in the Methods section of the initial publication. Of relevance to this work, all imaging data in the study represents T1-weighted structural brain scans that were processed using the Desikan-Killiany (DK) cortical atlas in Freesurfer^33^. They used univariate linear regression to identify group differences in the thickness and cortical area of the measured DK-atlas regions. Random-effects meta-analyses of both Cohen’s *d* and the partial correlation effect sizes were performed for each DK atlas region. Multiple testing correction was performed using the false discovery rate with a threshold or 0.05. Additional details on confounding and covariate analyses were included in the original publication. Following multiple testing correction and meta-analysis, 33 hemisphere-specific cortical surface area and thickness measures showed statistically significant differences between SCZ cases and healthy controls. Of the NIDPs reported by the UKB, we matched all 33 of these according to Freesurfer parcellation of T1-weighted structural imaging using the DK atlas.

### Maximally discriminatory GReX threshold determination

In the style of polygenic risk score analyses, we assessed the GReX-NIDP significance threshold at which the data in NeuroimaGene was maximally predictive of SCZ risk. We identified all NeuroimaGene GReX associations corresponding to the NIDPs and GReX from the SCZ studies carried out by the ENIGMA consortium and Trubetskoy et al. respectively. These represent associations between GReX measures and NIDPs that have each been associated with SCZ in independent studies. Beginning with a nominal p-value threshold of 0.05, we assessed the total number of associations that satisfied the threshold as well as the number of unique NIDPs represented by the association set. These represent two observed statistics: the number of SCZ GReX-NIDP associations that surpass the significance threshold and the number of unique NIDPs in the SCZ GReX-NIDP associations that surpass the significance threshold.

We then performed permutation testing to establish null distributions against which these statistics can be tested. To generate the null distribution, we selected 50 random genes (matching by 50 SCZ GReX measures that entered the experimental analysis). We then identified all NeuroimaGene associations that involve any of the 50 GReX measures and SCZ NIDPs at a p-value less than or equal to 0.05. Similarly to the experimental gene set, we then calculated null statistics detailing the total number of associations and the total number of unique NIDPs. We thus quantified the number of associations between SCZ NIDPs and *random* gene sets for comparison with associations between SCZ NIDP associations and *SCZ* GReX. We repeated this procedure 1000 times to estimate the distribution for each of the two test statistics “under the null.”

Using each empirically-derived distribution, we then identified the value below which 95% of null statistics fall. We performed this same analysis for the total number of GReX-NIDP associations and for the unique NIDPs included in each set, in line with the different null statistics (Figure 6b, Supplementary Figure 35a).

We repeated the permutation analysis at 6 different p-value thresholds to identify the threshold at which the association between SCZ GReX and SCZ NIDPs was maximally greater than for random genes. We show the difference between the experimental gene set statistics and the 95% threshold in Figure (6c) and Supplementary Figure 35b. From these data, we selected 0.005 as the optimal discriminatory threshold. Using this threshold, the value of the experimental test statistic relative to the simulated null distribution annotated with the 95^th^ percentile for each statistic is shown in Figure 6d for total GReX associations and Supplementary Figure 35c for unique NIDPs.

### Mendelian Randomization Joint Tissue Imputation

TWAS associations are susceptible to confounding from LD contamination. As a Mendelian Randomization approach, MR-JTI models variant-level heterogeneity to obtain an unbiased estimate the gene causal effect on phenotype. Using MR-JTI, we performed causal inference for the SCZ GReX associations.

We used the LiftOver script provided by the UCSC genome browser to convert the UKBB GWAS variant identifiers from Human Genome build 19 to build 38^34^. We then retrieved the full set of eQTL associations from the GTEx consortium Google cloud repository^31^. We intersected the retrieved eQTL data with each of the GWAS files from the UKB NIDPs prioritized by the initial TWAS. We harmonized SNPs between the two studies according to the chromosomal location and the effect/reference allele. We retrieved the LD scores obtained from the 1000 genomes project based on the RSID^35^. Using the MR-JTI script, we conducted Mendelian Randomization causal inference on these findings (See Code Availability).

## Data availability

The Neuroimaging TWAS resource is available on Zenodo at (to be provided upon publication). The summary statistics for the UKBB Neuroimaging GWAS are available at their BIG40 online data repository (https://open.win.ox.ac.uk/ukbiobank/big40/). Summary statistics for the ENIGMA dataset are available upon request from their website (https://enigma.ini.usc.edu/research/download-enigma-gwas-results/). Processed GTEx data (for example, gene expression and eQTLs) are available from the GTEx portal (https://gtexportal.org). The Chain files required for running the UCSC LiftOver script are accessible upon request from the UCSC genome store (https://genome-store.ucsc.edu/). LD scores from the 1000 genomes project were downloaded from the google repository maintained by the Alkes Price group at the Broad Institute (https://alkesgroup.broadinstitute.org/LDSCORE/) in the file labeled “1000G_Phase3_ldscores.tgz”. The tissue specific gene expression models trained on GTEx and enriched via JTI are available for download from Zenodo (https://doi.org/10.5281/zenodo.3842289).

## Code availability

The command line script required to run the UCSC LiftOver tool is available upon request from the UCSC genome store (https://genome-store.ucsc.edu/). Code for MultiXcan is available on the Hakyim Github repository (https://github.com/hakyimlab/MetaXcan). Code for the JTI model training and MR-JTI causal inference analysis is hosted at the Gamazon Github repository (https://github.com/gamazonlab/MR-JTI). All code required for the reproduction of this analysis and replication of all tables and figures is hosted at the corresponding author’s Github repository (https://github.com/xbledsoe/NeuroimaGene). Data required for replication as well as large supplementary data sets can be accessed at (https://zenodo.org).

**[these last two resource links will be made active upon publication.]

## Data Availability

The Neuroimaging TWAS resource is available on Zenodo at (to be provided upon publication). The summary statistics for the UKBB Neuroimaging GWAS are available at their BIG40 online data repository (https://open.win.ox.ac.uk/ukbiobank/big40/). Summary Statistics for the replication dataset in ENIGMA are available upon request from their website (https://enigma.ini.usc.edu/research/download-enigma-gwas-results/). Processed GTEx data (for example, gene expression and eQTLs) are available from the GTEx portal (https://gtexportal.org). The Chain files required for running the UCSC LiftOver script are accessible upon request from the UCSC genome store (https://genome-store.ucsc.edu/). LD scores from the 1000 genomes project are available for download from the google repository maintained by the Alkes group at the Broad Institute (https://alkesgroup.broadinstitute.org/LDSCORE/) in the file labeled 1000G_Phase3_ldscores.tgz. The tissue specific gene expression models trained on GTEx and enriched via JTI are available for download from Zenodo (https://doi.org/10.5281/zenodo.3842289).

## Acknowledgements

We acknowledge support from the following National Institutes of Health (NIH) grants (to E.R.G.): R35HG010718, NHGRI R01HG011138, NIMH R01MH126459, NIGMS R01GM140287, and NIA AG068026. We thank Vanderbilt’s Advanced Computing Center for Research and Education (ACCRE) for infrastructure support.

