## Supplementary Figures for "A Transcriptomic Atlas of the Human Brain Reveals Genetically Determined Aspects of Neuropsychiatric Health"

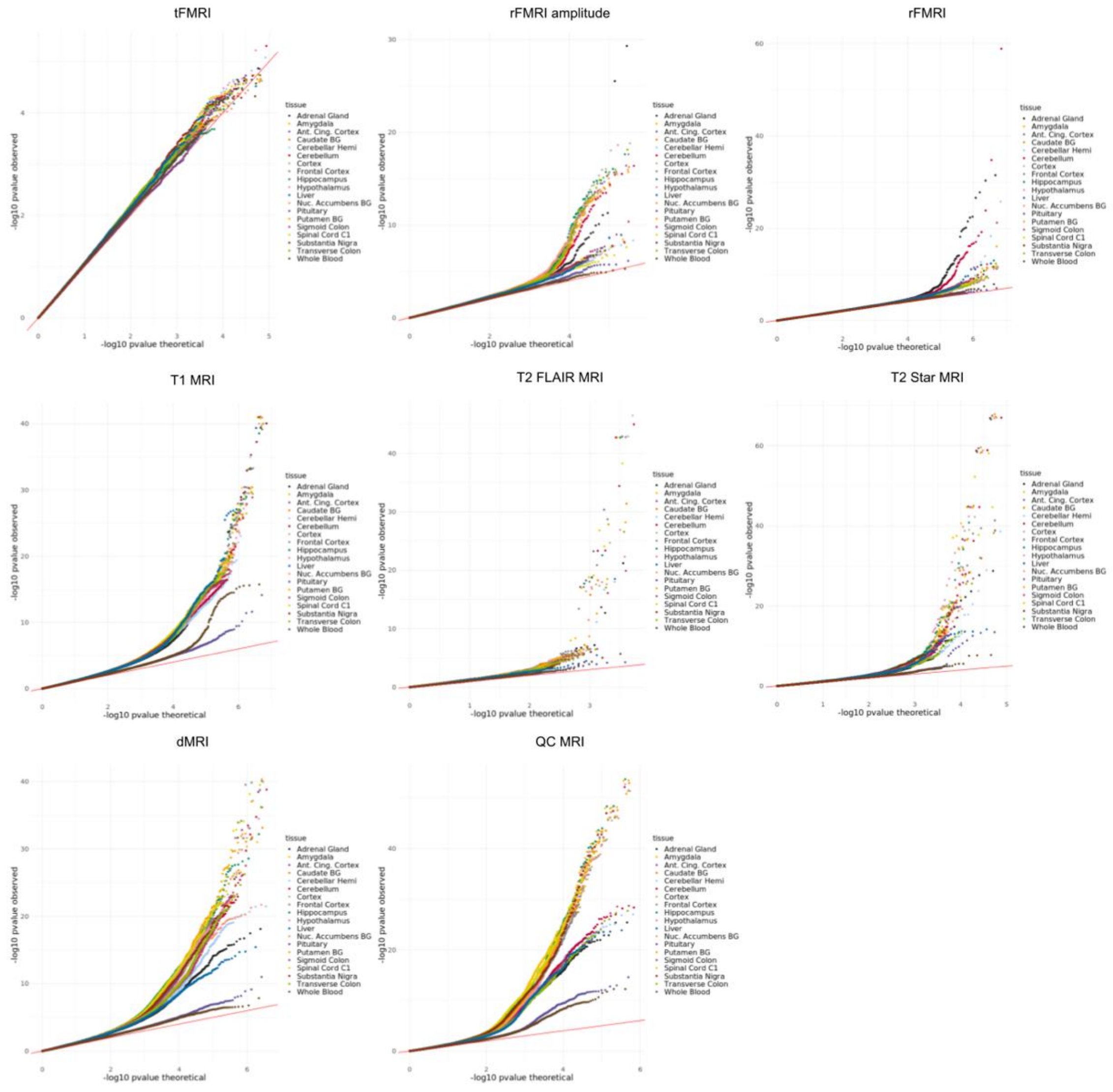


**Supplementary Figure 1:** Quantile-quantile plots of all Neuroimaging TWAS associations paneled by MR imaging modality and colored according to JTI tissue model.

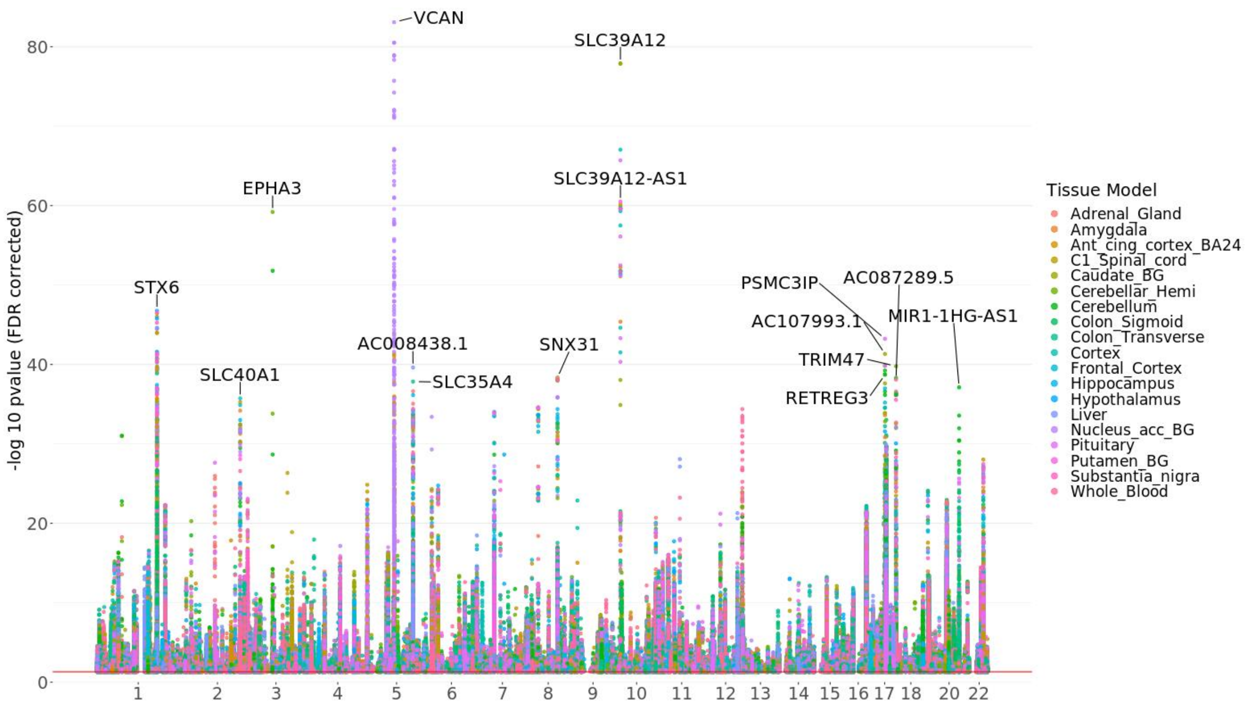


**Supplementary Figure 2**: Manhattan style plot of all GReX associations with NIDPs across the genome according to all 19 JTI tissue models including findings from the inversion regions at Chr17 and Chr8, the extended MHC region, and GReX associations with predicted performance correlations of <0.1.

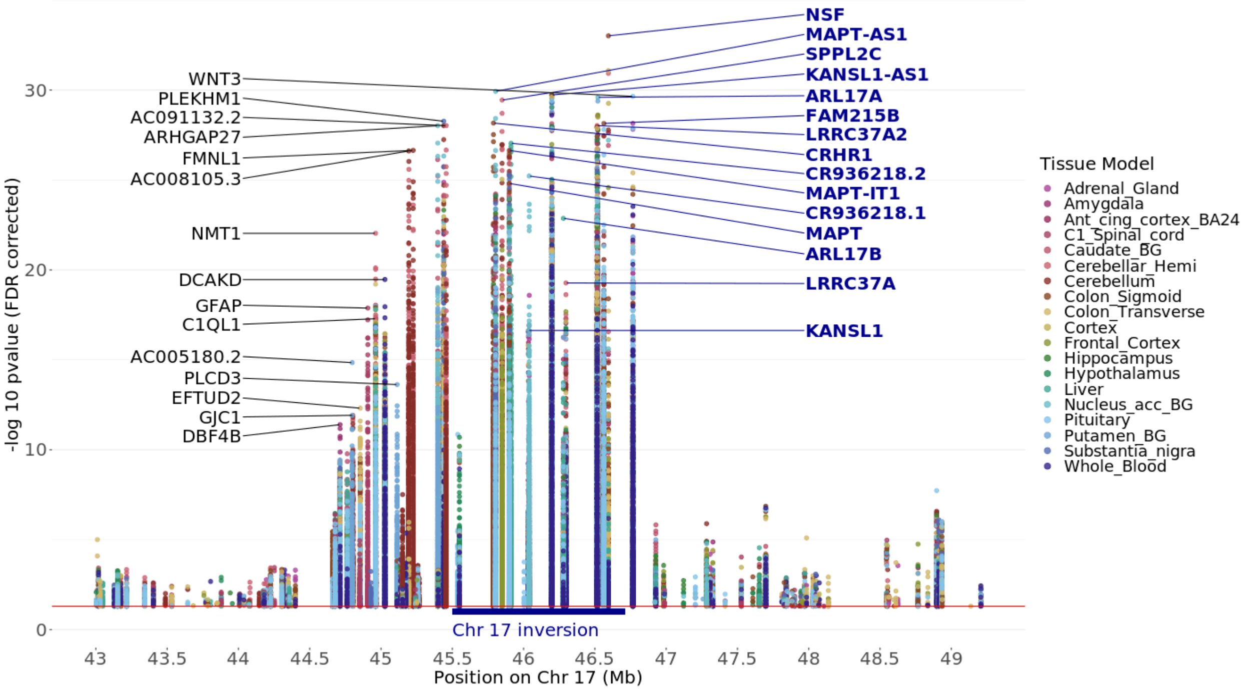


**Supplementary Figure 3**: Manhattan style plot of GReX associations with NIDPs at the 17q21.3 locus including the MAPT inversion (chr17: 45495836 – 46707123)[1] according to all 19 JTI tissue models including GReX associations with predicted performance correlations of <0.1. Flanking regions of 1MB are included for annotation and the top 30 genes are listed.

[1] Campoy, Elena, et al. "Genomic architecture and functional effects of potential human inversion supergenes." Philosophical Transactions of the Royal Society B 377.1856 (2022): 20210209.

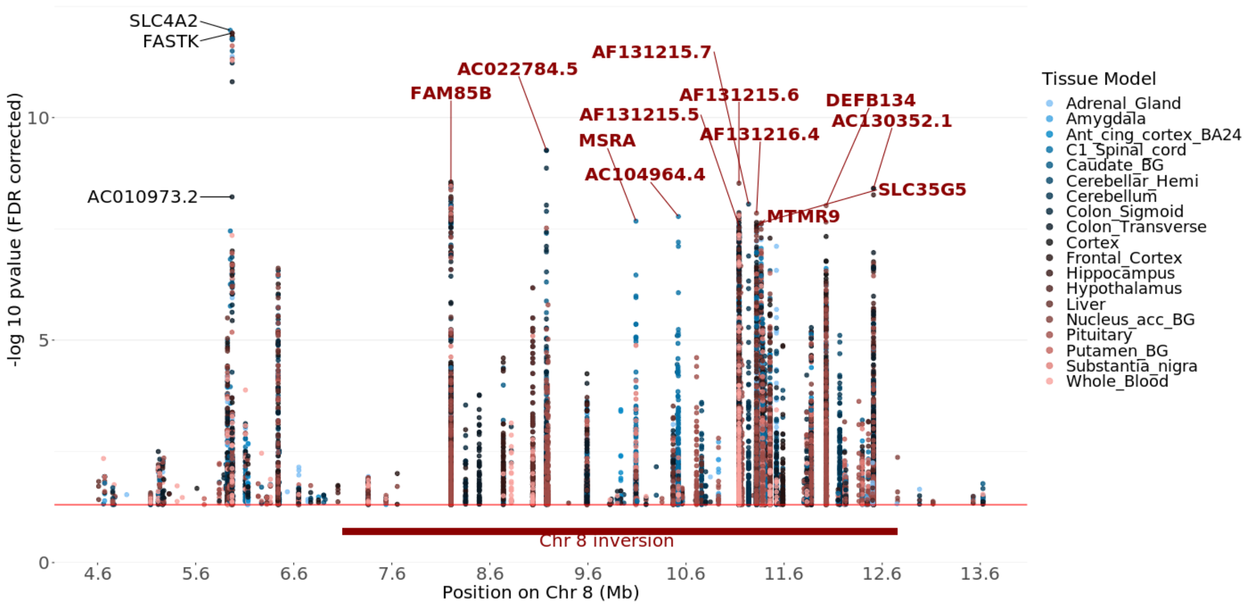


**Supplementary Figure 4**: Manhattan style plot of GReX associations with NIDPs at the 8p23.1 locus (chr8: 7064966 - 7064966)[1] with 1MB flanking regions according to all 19 JTI tissue models including GReX associations with predicted performance correlations of <0.1. The top 15 genes are listed.

[1] Campoy, Elena, et al. "Genomic architecture and functional effects of potential human inversion supergenes." Philosophical Transactions of the Royal Society B 377.1856 (2022): 20210209.

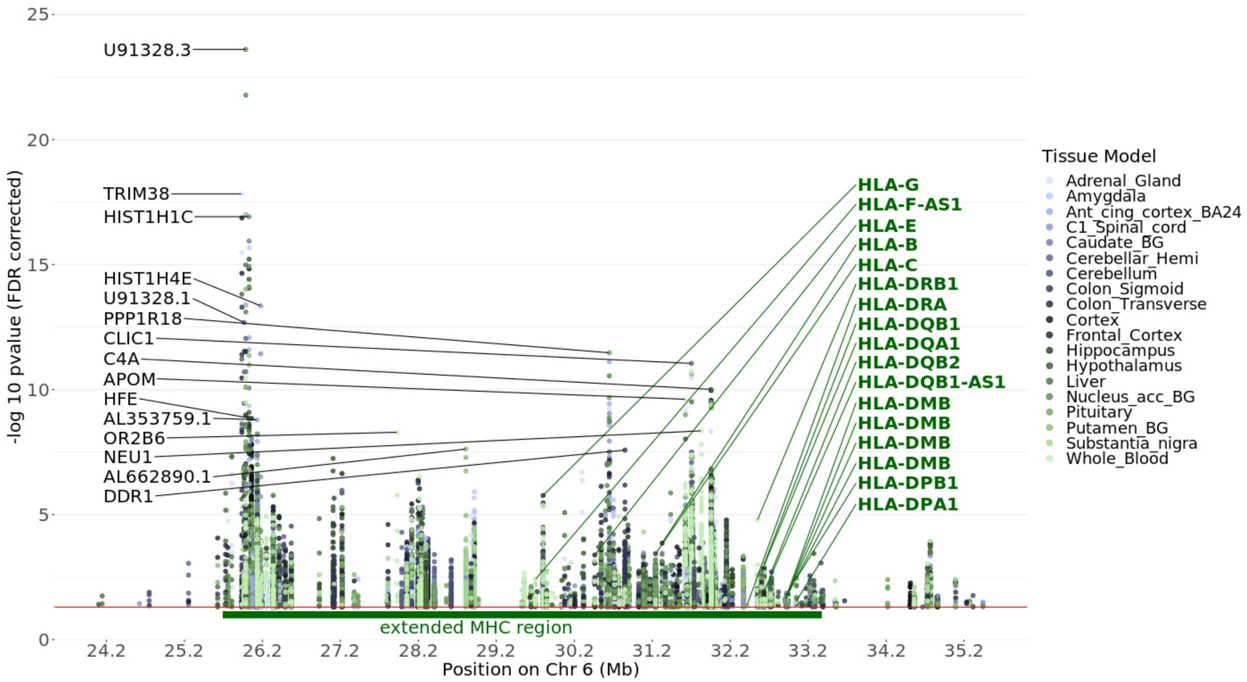


**Supplementary Figure 5**: Manhattan style plot of GReX associations with NIDPs at the extended MHC region (chr6: 25726063 - 33400644)[1] according to all 19 JTI tissue models including GReX associations with predicted performance correlations of <0.1. The top 30 genes are listed.

[1] Horton, R., Wilming, L., Rand, V. et al. Gene map of the extended human MHC. Nat Rev Genet 5, 889–899 (2004).

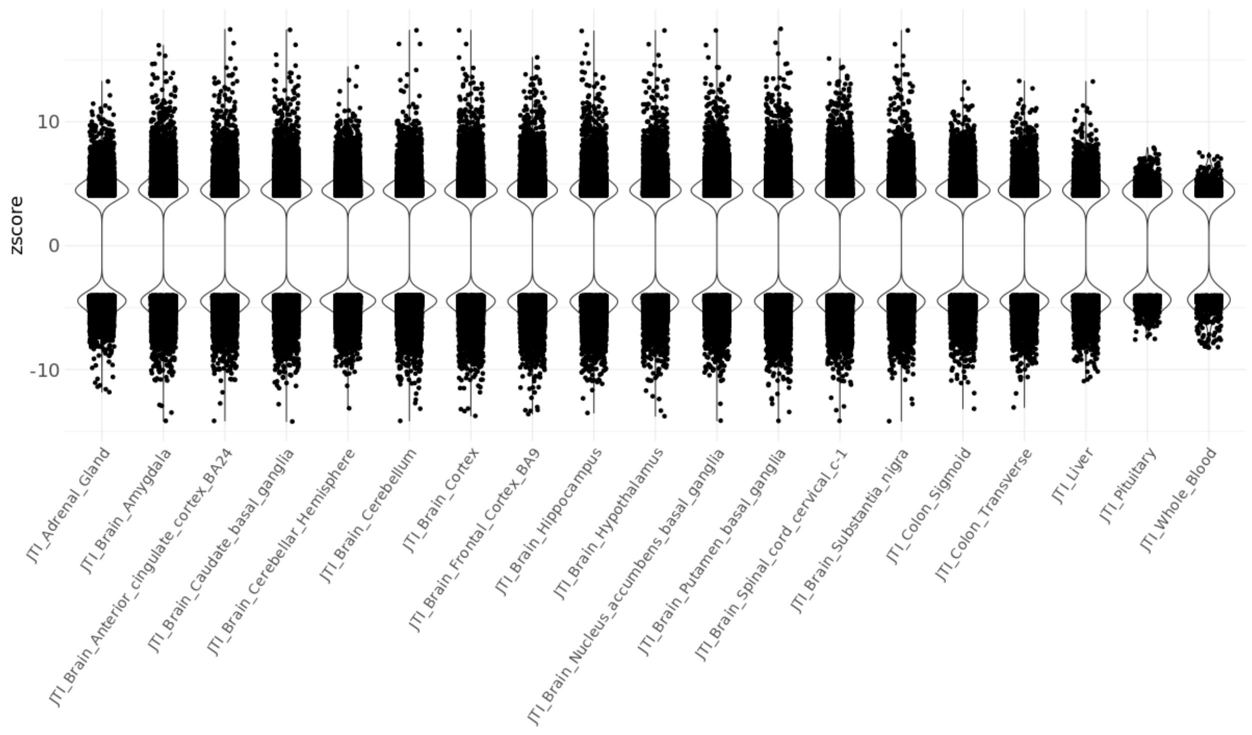


**Supplementary Figure 6:** Distribution of normalized effect sizes for associations in each JTI-enriched eQTL model. This visualization represents the findings that surpass the study-wide FDR threshold.

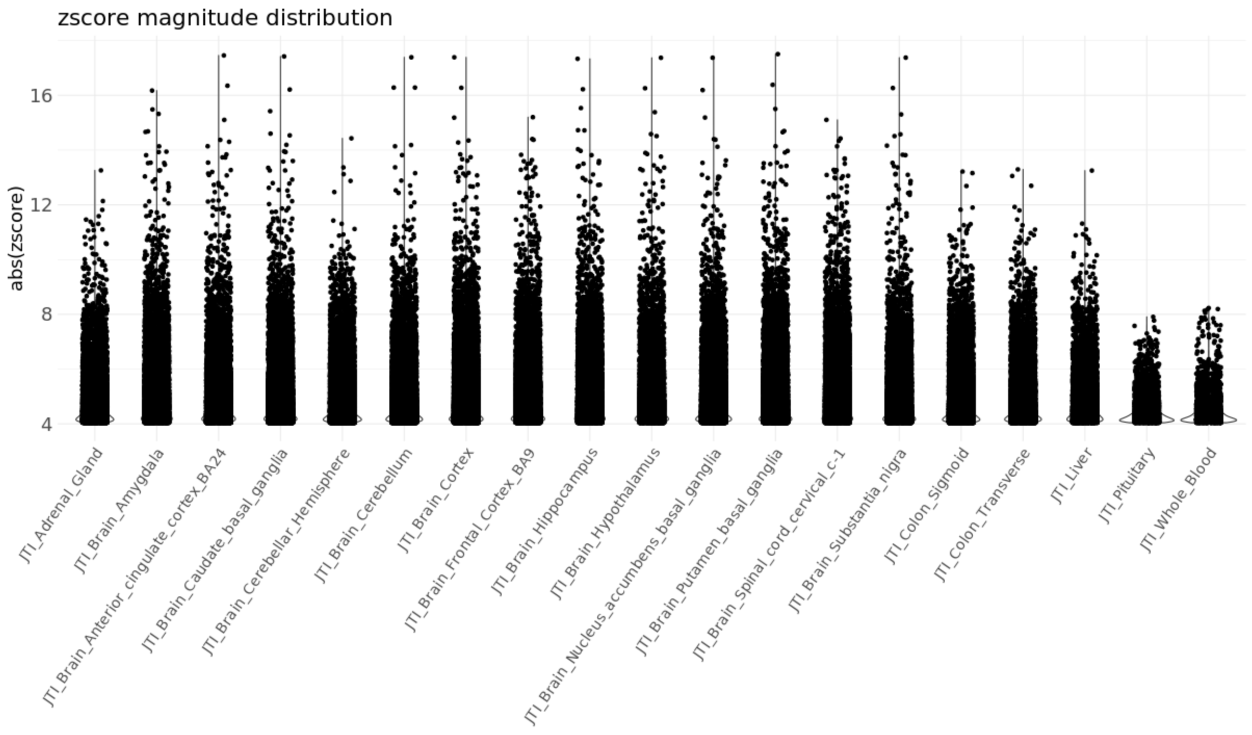


**Supplementary Figure 7**: Distribution of the magnitude of normalized effect sizes for associations in each JTI-enriched eQTL model. This visualization represents the findings that surpass the study-wide FDR threshold.

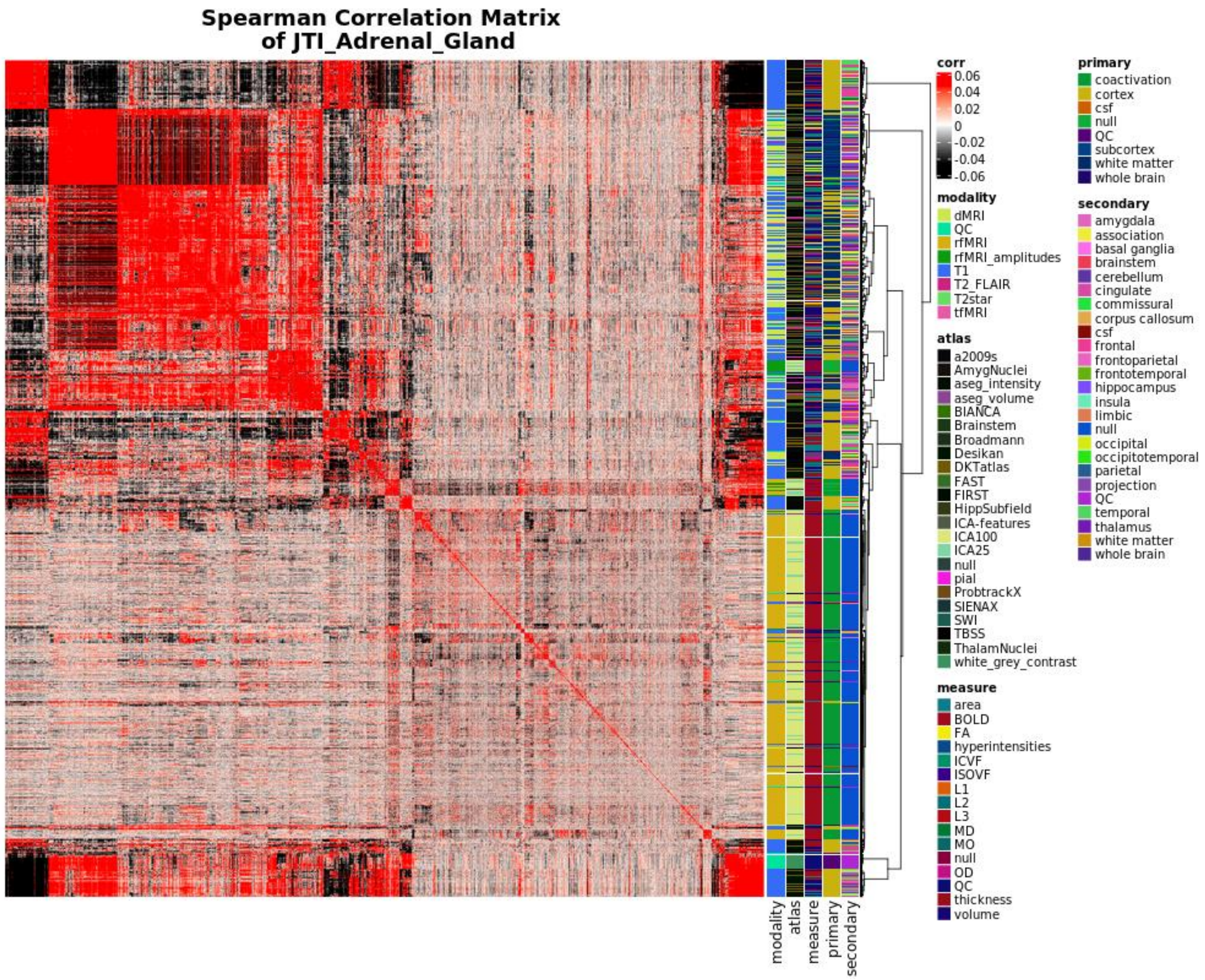


**Supplementary Figure 8:** We demonstrate NIPD similarity on the basis of GReX association effect size with a matrix detailing the pairwise spearman correlation distance between all NIDPs according to the unfiltered z-scores of all measured GReX associations as predicted by the adrenal gland. Annotations describe the MR imaging modality (modality), the atlas used in the extraction of pre-defined neurologic regions (atlas), the type of measurement characterized by each NIDP (measure), the type of brain tissue/MR element being examined (primary), and the named region of the brain characterized by the NIDP (secondary). These values are derived from the UKB supplementary NIDP information and detailed in Supplementary Table 1. Corresponding ranked ordering of NIDPs according to this tissue model can be found in Supplementary Table 2.

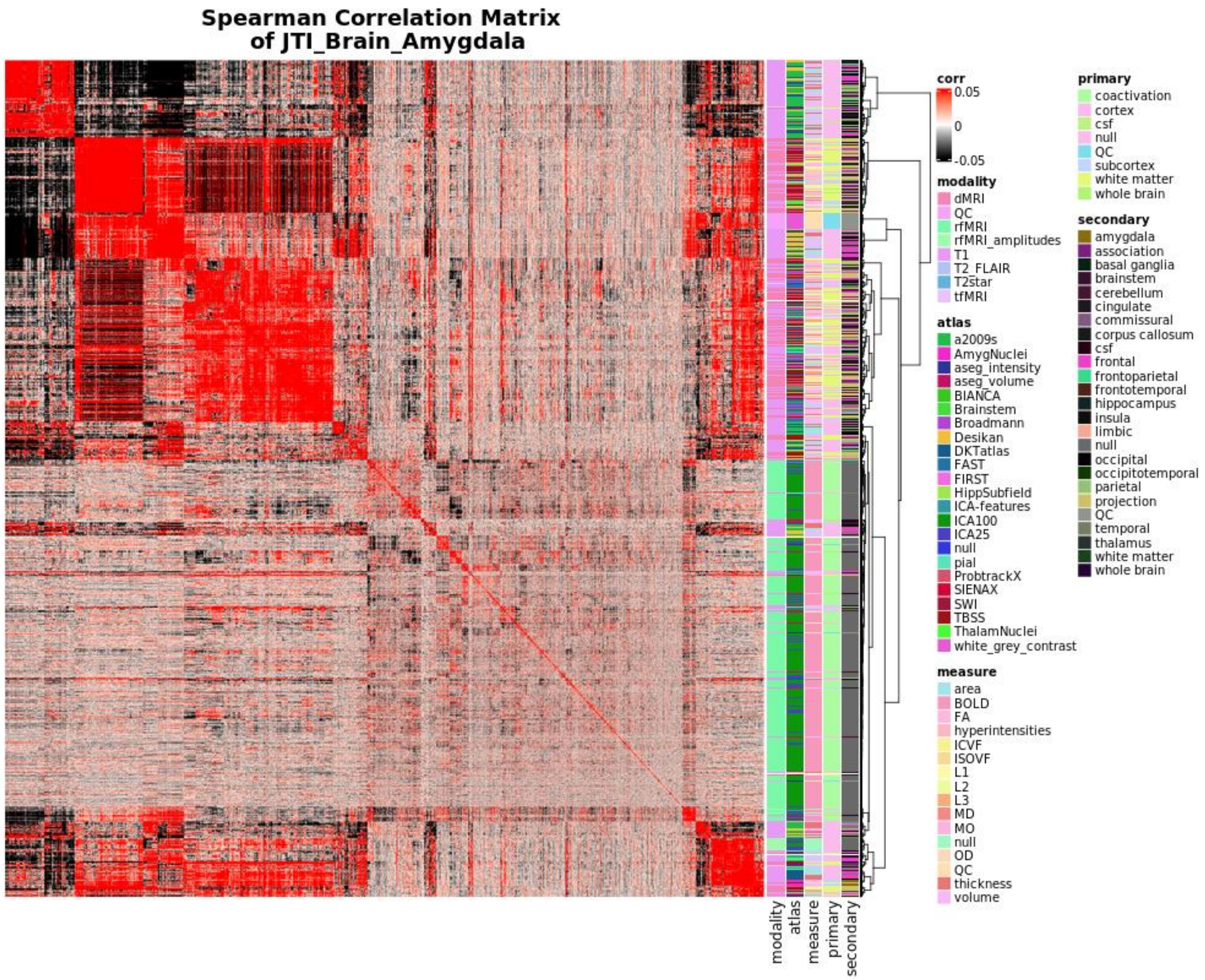


**Supplementary Figure 9:** We demonstrate NIPD similarity on the basis of GReX association effect size with a matrix detailing the pairwise spearman correlation distance between all NIDPs according to the unfiltered z-scores of all measured GReX associations as predicted by the amygdala. Annotations describe the MR imaging modality (modality), the atlas used in the extraction of pre-defined neurologic regions (atlas), the type of measurement characterized by each NIDP (measure), the type of brain tissue/MR element being examined (primary), and the named region of the brain characterized by the NIDP (secondary). These values are derived from the UKB supplementary NIDP information and detailed in Supplementary Table 1. Corresponding ranked ordering of NIDPs according to this tissue model can be found in Supplementary Table 3.

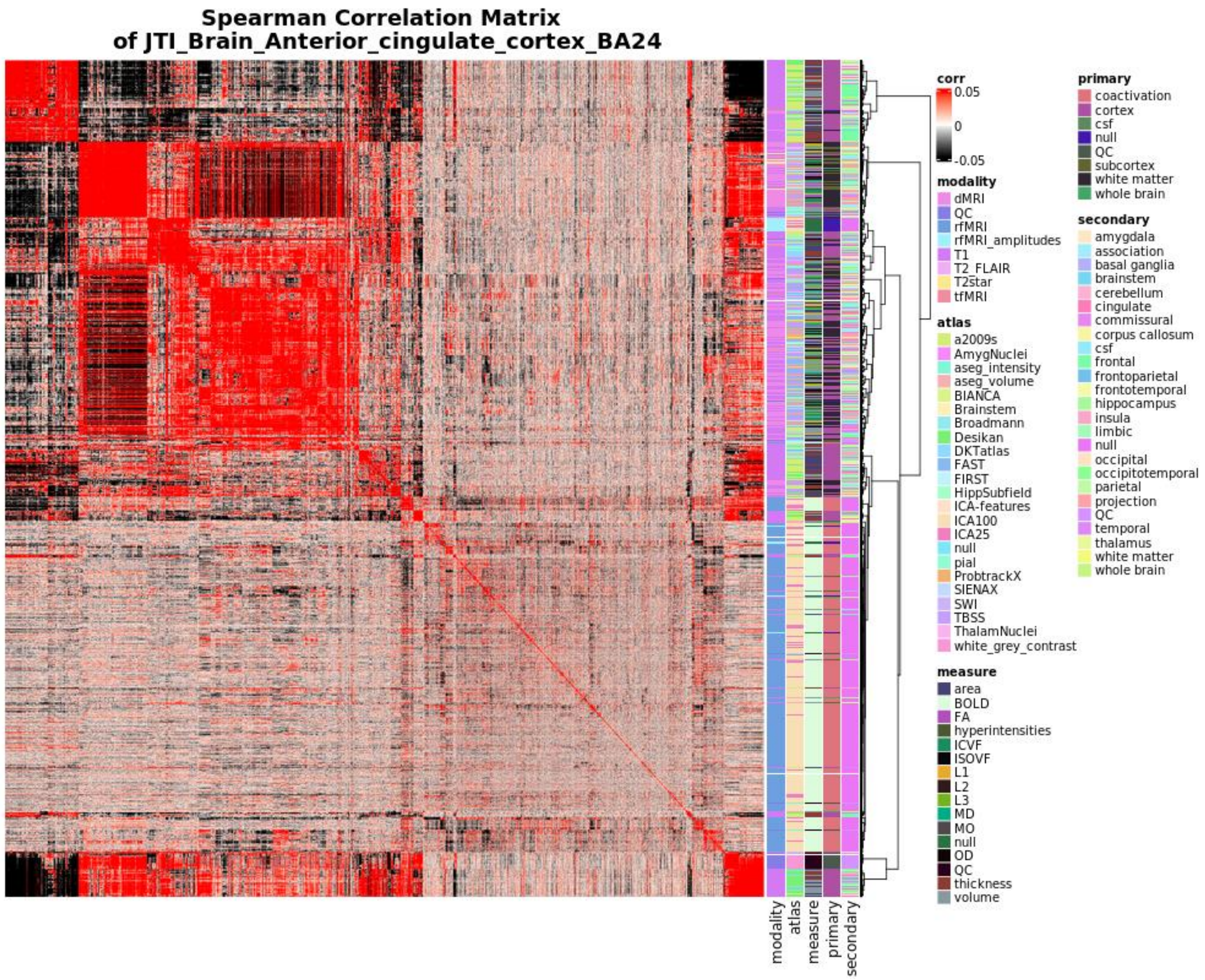


**Supplementary Figure 10:** We demonstrate NIPD similarity on the basis of GReX association effect size with a matrix detailing the pairwise spearman correlation distance between all NIDPs according to the unfiltered z-scores of all measured GReX associations as predicted by the anterior cingulate cortex. Annotations describe the MR imaging modality (modality), the atlas used in the extraction of pre-defined neurologic regions (atlas), the type of measurement characterized by each NIDP (measure), the type of brain tissue/MR element being examined (primary), and the named region of the brain characterized by the NIDP (secondary). These values are derived from the UKB supplementary NIDP information and detailed in Supplementary Table 1. Corresponding ranked ordering of NIDPs according to this tissue model can be found in Supplementary Table 4.

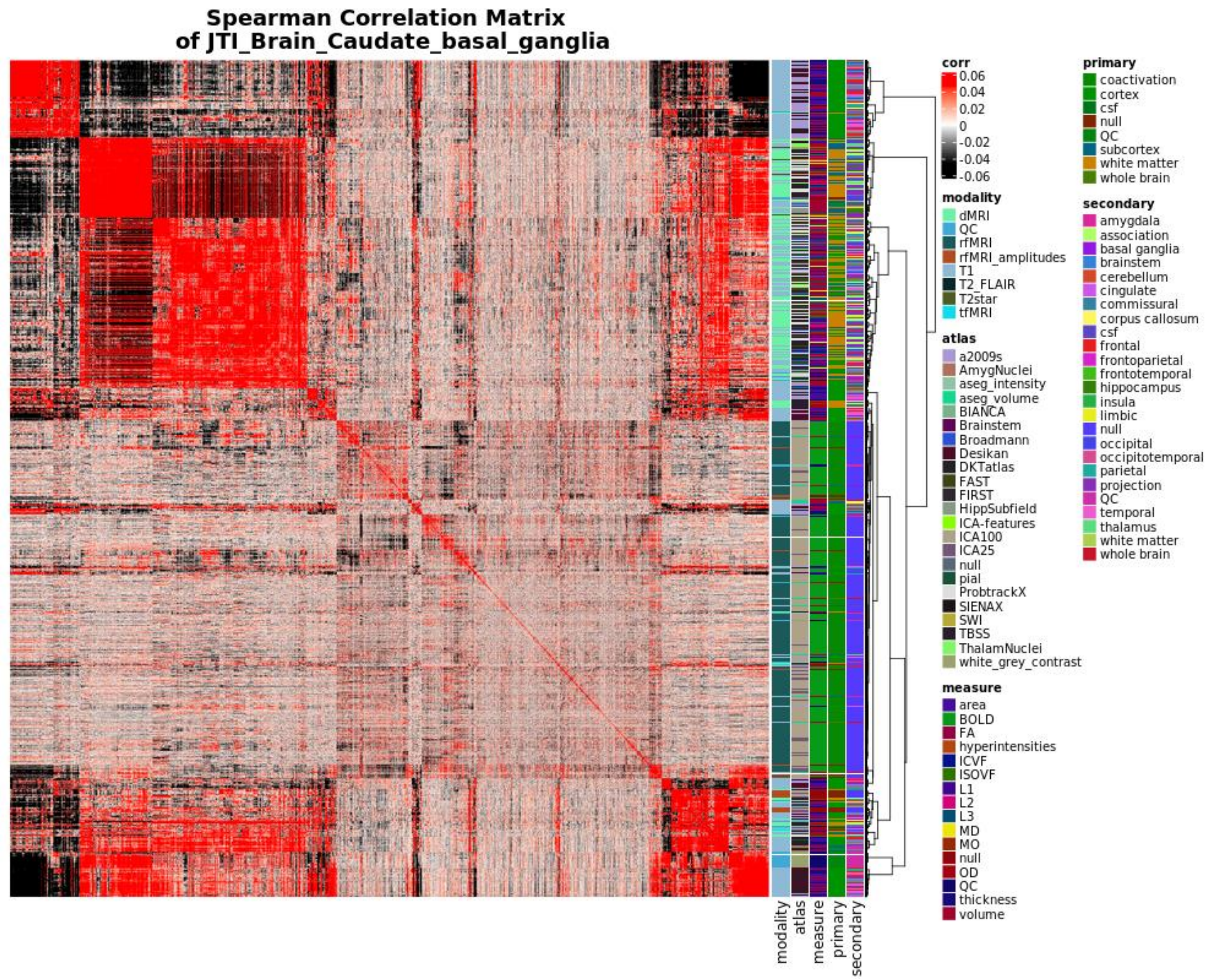


**Supplementary Figure 11:** We demonstrate NIPD similarity on the basis of GReX association effect size with a matrix detailing the pairwise spearman correlation distance between all NIDPs according to the unfiltered z-scores of all measured GReX associations as predicted by the caudate basal ganglia. Annotations describe the MR imaging modality (modality), the atlas used in the extraction of pre-defined neurologic regions (atlas), the type of measurement characterized by each NIDP (measure), the type of brain tissue/MR element being examined (primary), and the named region of the brain characterized by the NIDP (secondary). These values are derived from the UKB supplementary NIDP information and detailed in Supplementary Table 1. Corresponding ranked ordering of NIDPs according to this tissue model can be found in Supplementary Table 5.

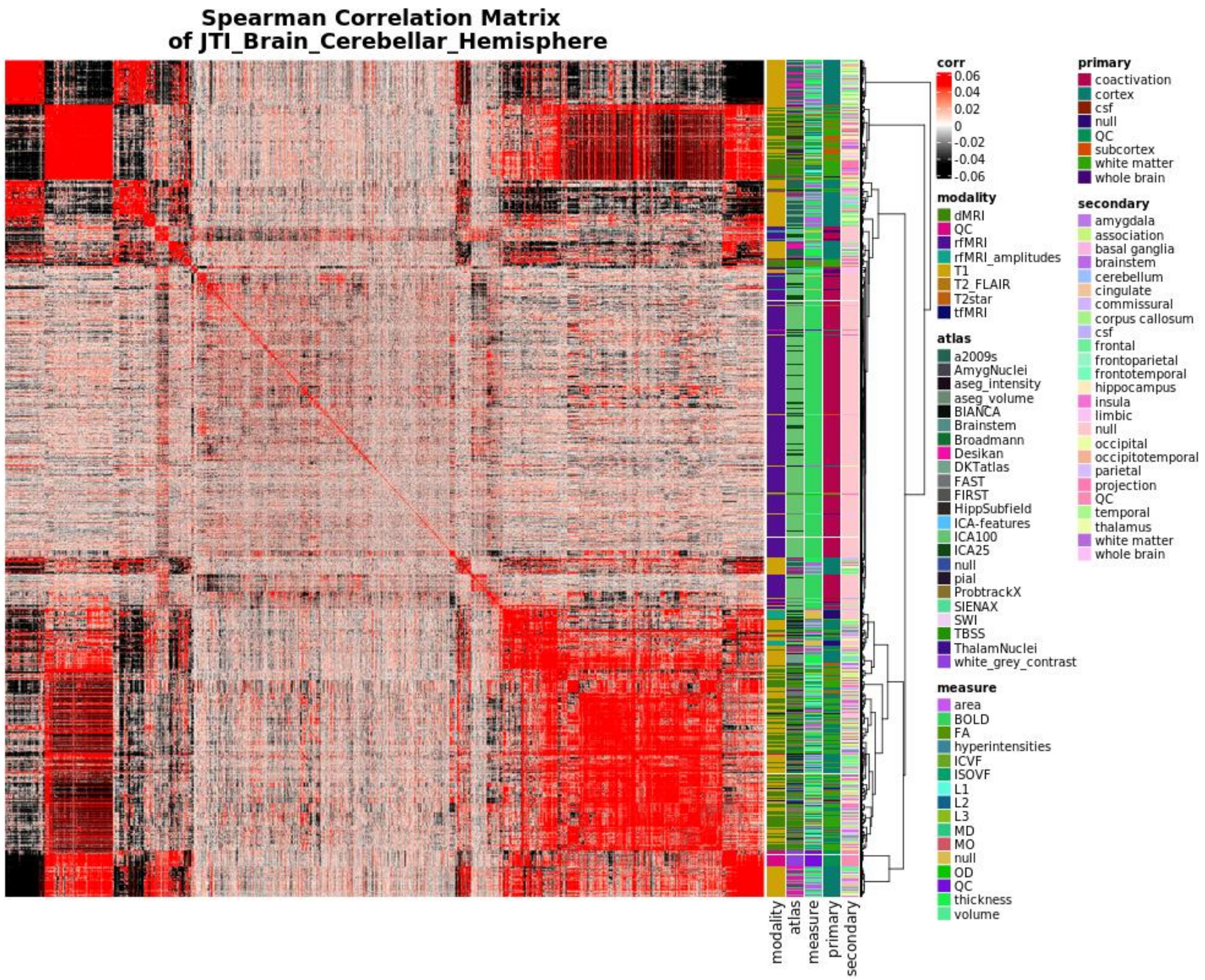


**Supplementary Figure 12:** We demonstrate NIPD similarity on the basis of GReX association effect size with a matrix detailing the pairwise spearman correlation distance between all NIDPs according to the unfiltered z-scores of all measured GReX associations as predicted by the cerebellar hemisphere. Annotations describe the MR imaging modality (modality), the atlas used in the extraction of pre-defined neurologic regions (atlas), the type of measurement characterized by each NIDP (measure), the type of brain tissue/MR element being examined (primary), and the named region of the brain characterized by the NIDP (secondary). These values are derived from the UKB supplementary NIDP information and detailed in Supplementary Table 1. Corresponding ranked ordering of NIDPs according to this tissue model can be found in Supplementary Table 6.

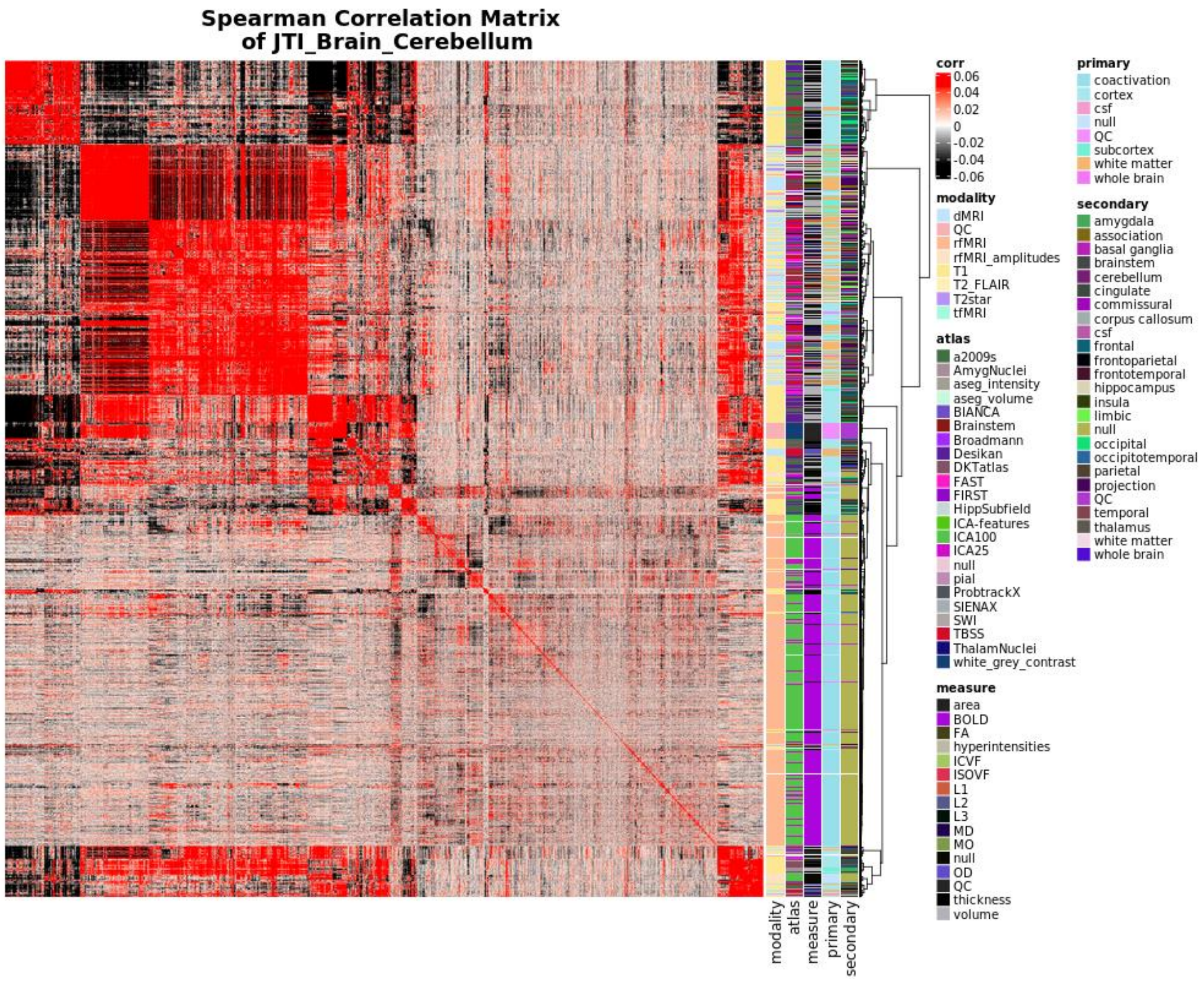


**Supplementary Figure 13:** We demonstrate NIPD similarity on the basis of GReX association effect size with a matrix detailing the pairwise spearman correlation distance between all NIDPs according to the unfiltered z-scores of all measured GReX associations as predicted by the cerebellum. Annotations describe the MR imaging modality (modality), the atlas used in the extraction of pre-defined neurologic regions (atlas), the type of measurement characterized by each NIDP (measure), the type of brain tissue/MR element being examined (primary), and the named region of the brain characterized by the NIDP (secondary). These values are derived from the UKB supplementary NIDP information and detailed in Supplementary Table 1. Corresponding ranked ordering of NIDPs according to this tissue model can be found in Supplementary Table 7.

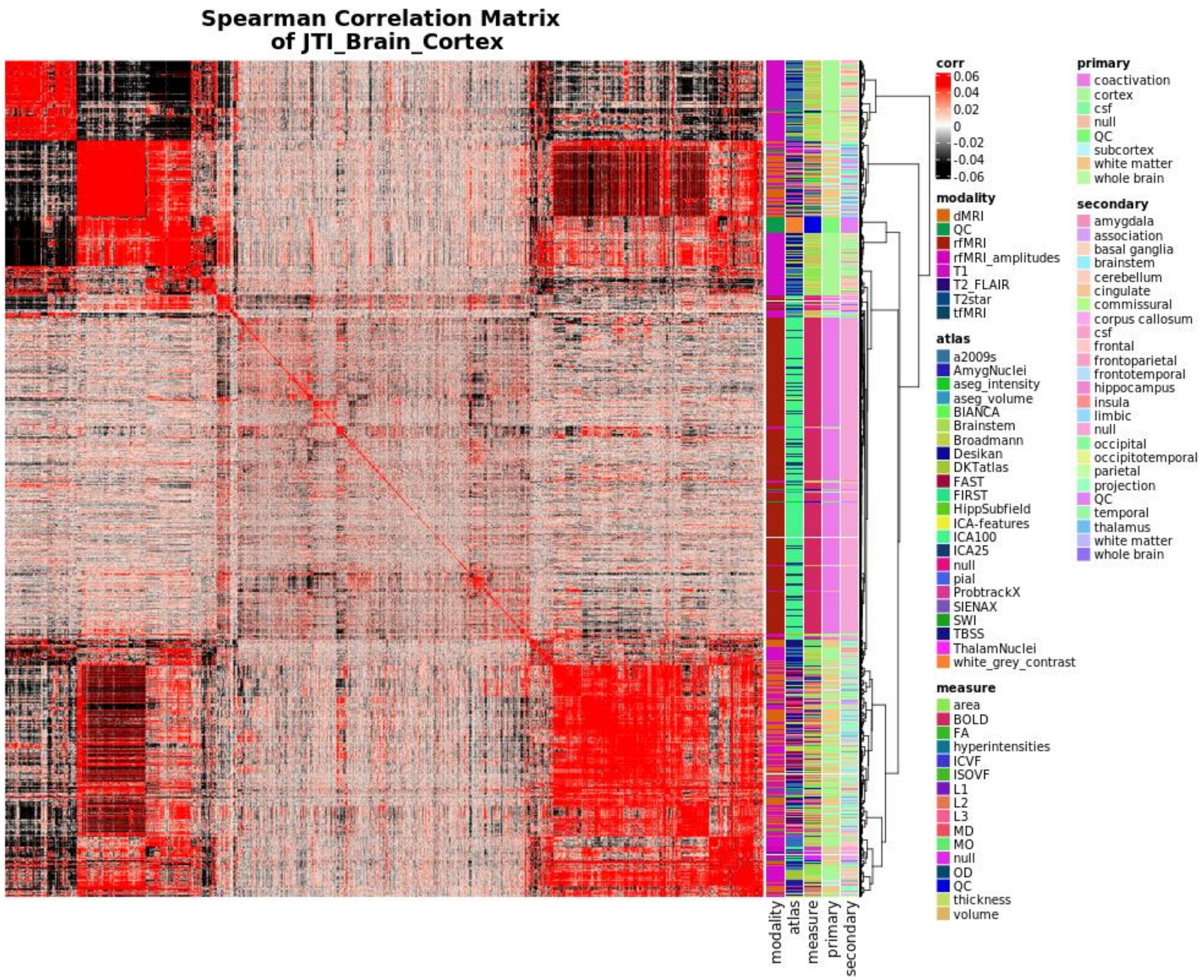


**Supplementary Figure 14:** We demonstrate NIPD similarity on the basis of GReX association effect size with a matrix detailing the pairwise spearman correlation distance between all NIDPs according to the unfiltered z-scores of all measured GReX associations as predicted by the cortex. Annotations describe the MR imaging modality (modality), the atlas used in the extraction of pre-defined neurologic regions (atlas), the type of measurement characterized by each NIDP (measure), the type of brain tissue/MR element being examined (primary), and the named region of the brain characterized by the NIDP (secondary). These values are derived from the UKB supplementary NIDP information and detailed in Supplementary Table 1. Corresponding ranked ordering of NIDPs according to this tissue model can be found in Supplementary Table 8.

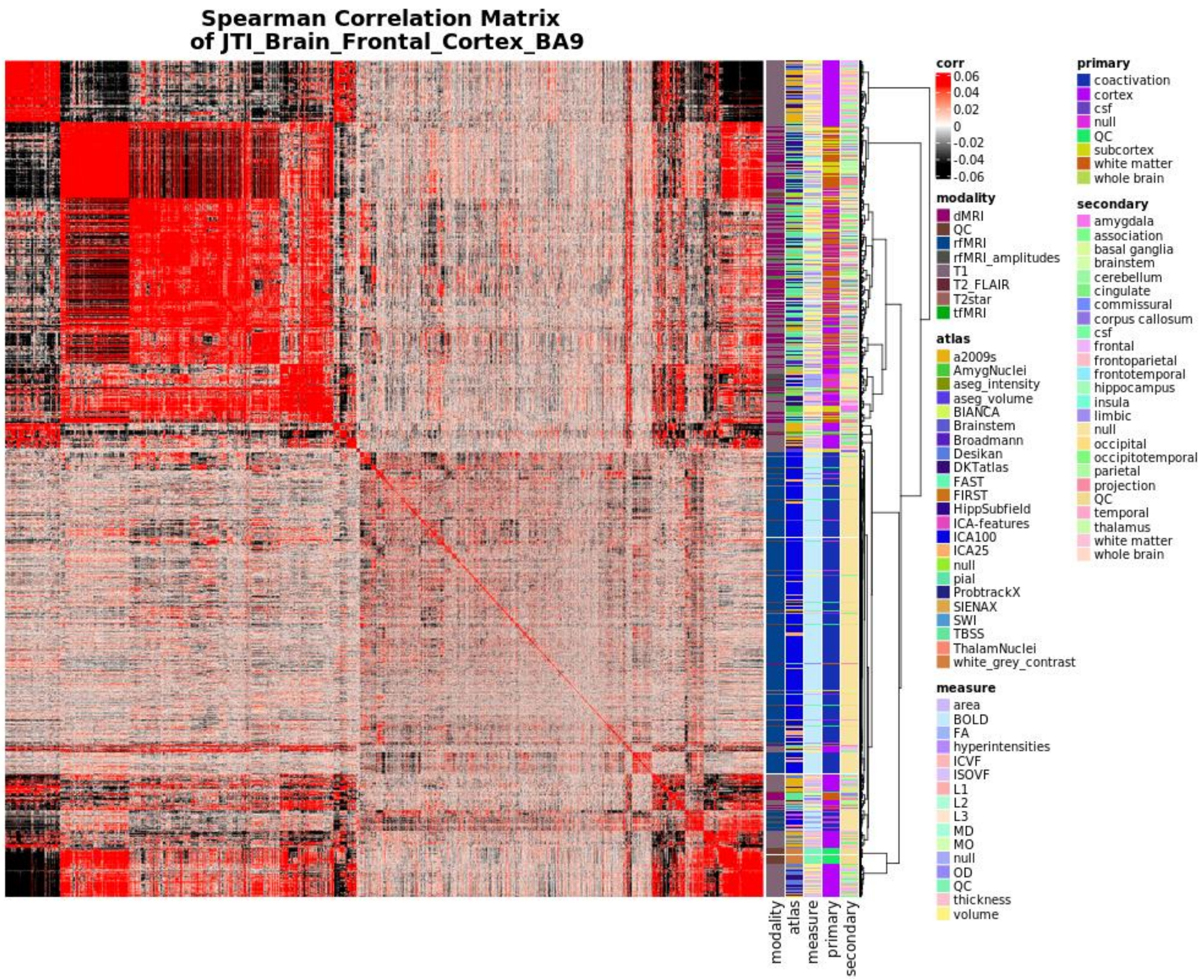


**Supplementary Figure 15:** We demonstrate NIPD similarity on the basis of GReX association effect size with a matrix detailing the pairwise spearman correlation distance between all NIDPs according to the unfiltered z-scores of all measured GReX associations as predicted by the frontal cortex. Annotations describe the MR imaging modality (modality), the atlas used in the extraction of pre-defined neurologic regions (atlas), the type of measurement characterized by each NIDP (measure), the type of brain tissue/MR element being examined (primary), and the named region of the brain characterized by the NIDP (secondary). These values are derived from the UKB supplementary NIDP information and detailed in Supplementary Table 1. Corresponding ranked ordering of NIDPs according to this tissue model can be found in Supplementary Table 9.

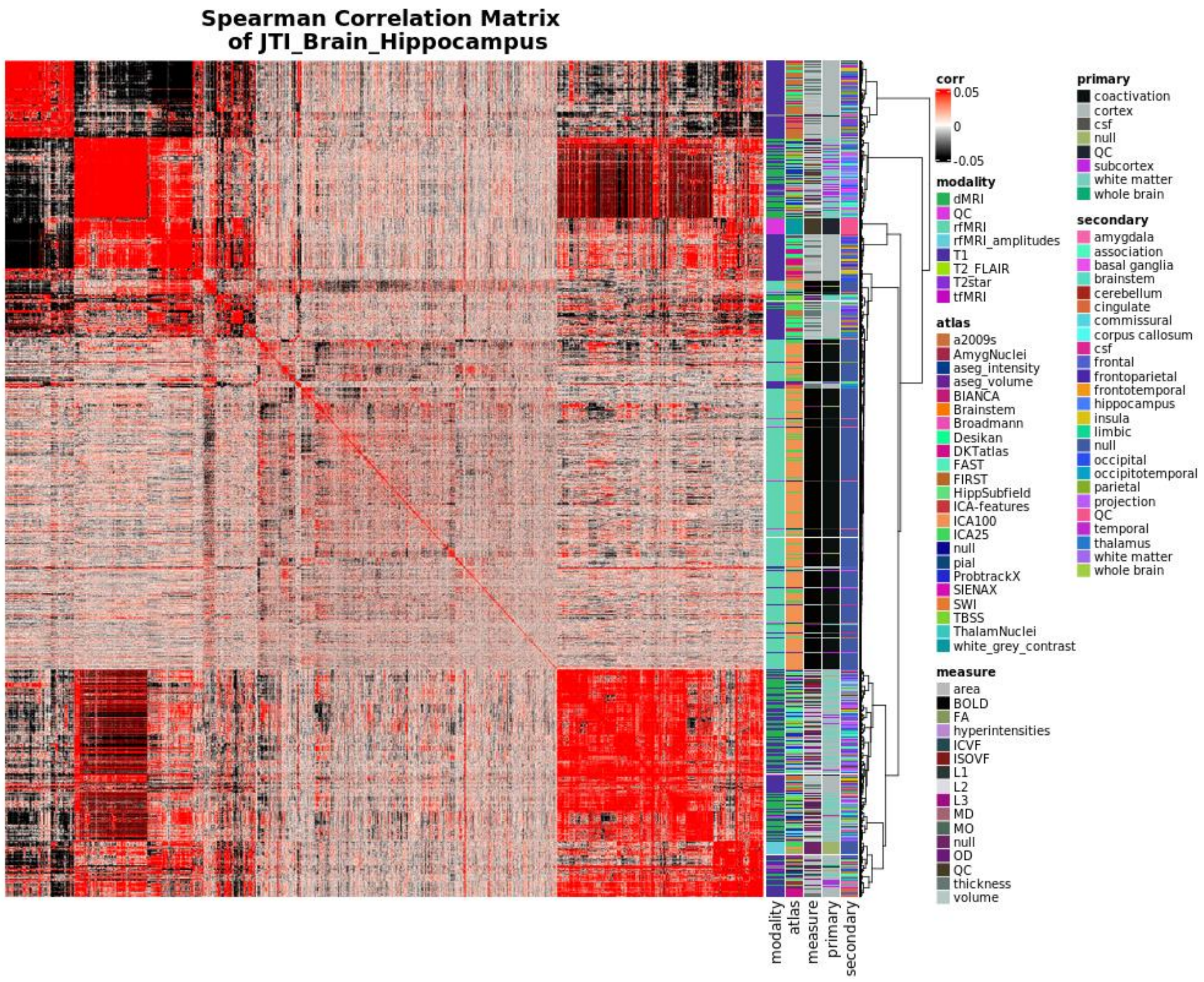


**Supplementary Figure 16:** We demonstrate NIPD similarity on the basis of GReX association effect size with a matrix detailing the pairwise spearman correlation distance between all NIDPs according to the unfiltered z-scores of all measured GReX associations as predicted by the hippocampus. Annotations describe the MR imaging modality (modality), the atlas used in the extraction of pre-defined neurologic regions (atlas), the type of measurement characterized by each NIDP (measure), the type of brain tissue/MR element being examined (primary), and the named region of the brain characterized by the NIDP (secondary). These values are derived from the UKB supplementary NIDP information and detailed in Supplementary Table 1. Corresponding ranked ordering of NIDPs according to this tissue model can be found in Supplementary Table 10.

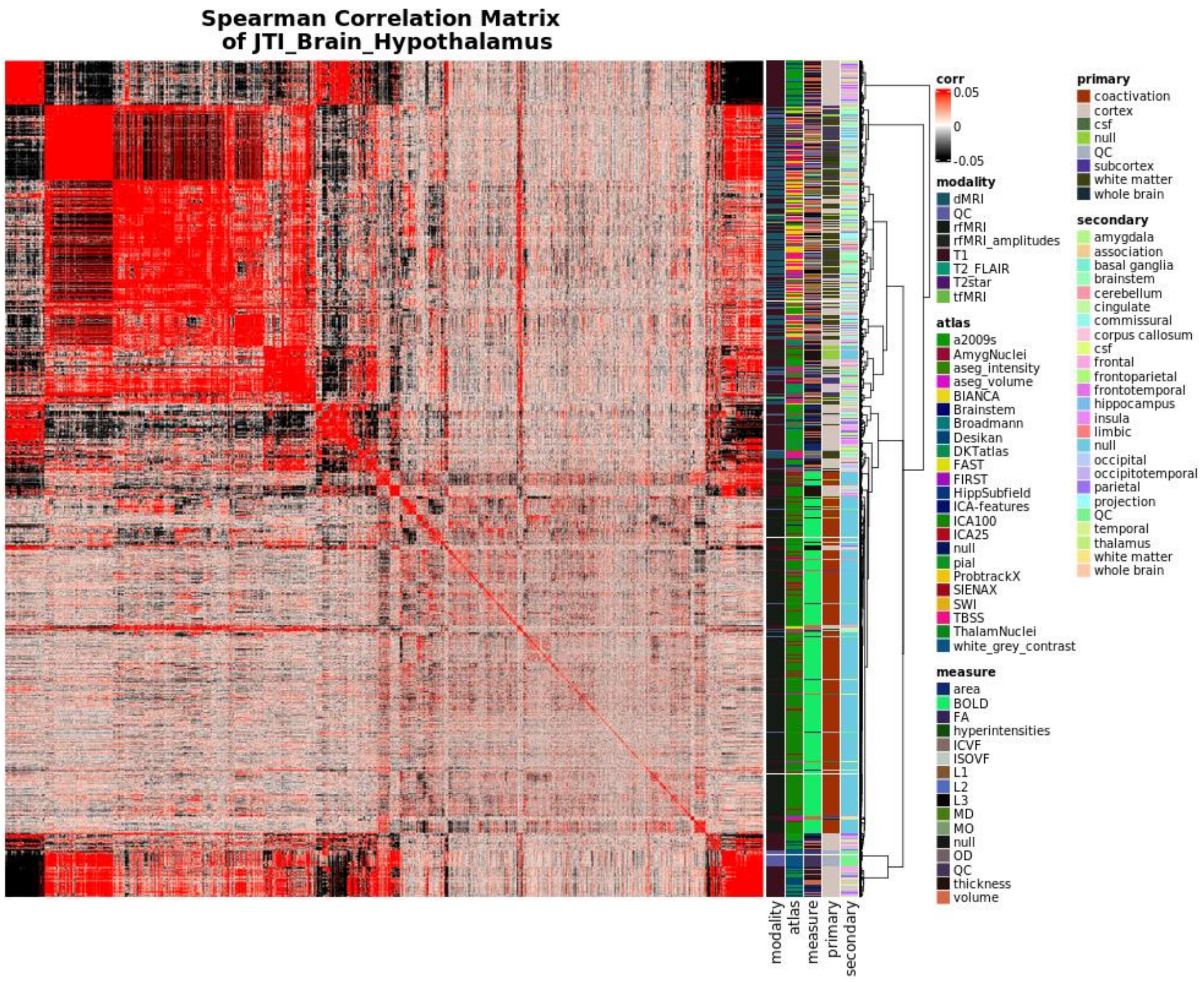


**Supplementary Figure 17:** We demonstrate NIPD similarity on the basis of GReX association effect size with a matrix detailing the pairwise spearman correlation distance between all NIDPs according to the unfiltered z-scores of all measured GReX associations as predicted by the hypothalamus. Annotations describe the MR imaging modality (modality), the atlas used in the extraction of pre-defined neurologic regions (atlas), the type of measurement characterized by each NIDP (measure), the type of brain tissue/MR element being examined (primary), and the named region of the brain characterized by the NIDP (secondary). These values are derived from the UKB supplementary NIDP information and detailed in Supplementary Table 1. Corresponding ranked ordering of NIDPs according to this tissue model can be found in Supplementary Table 11.

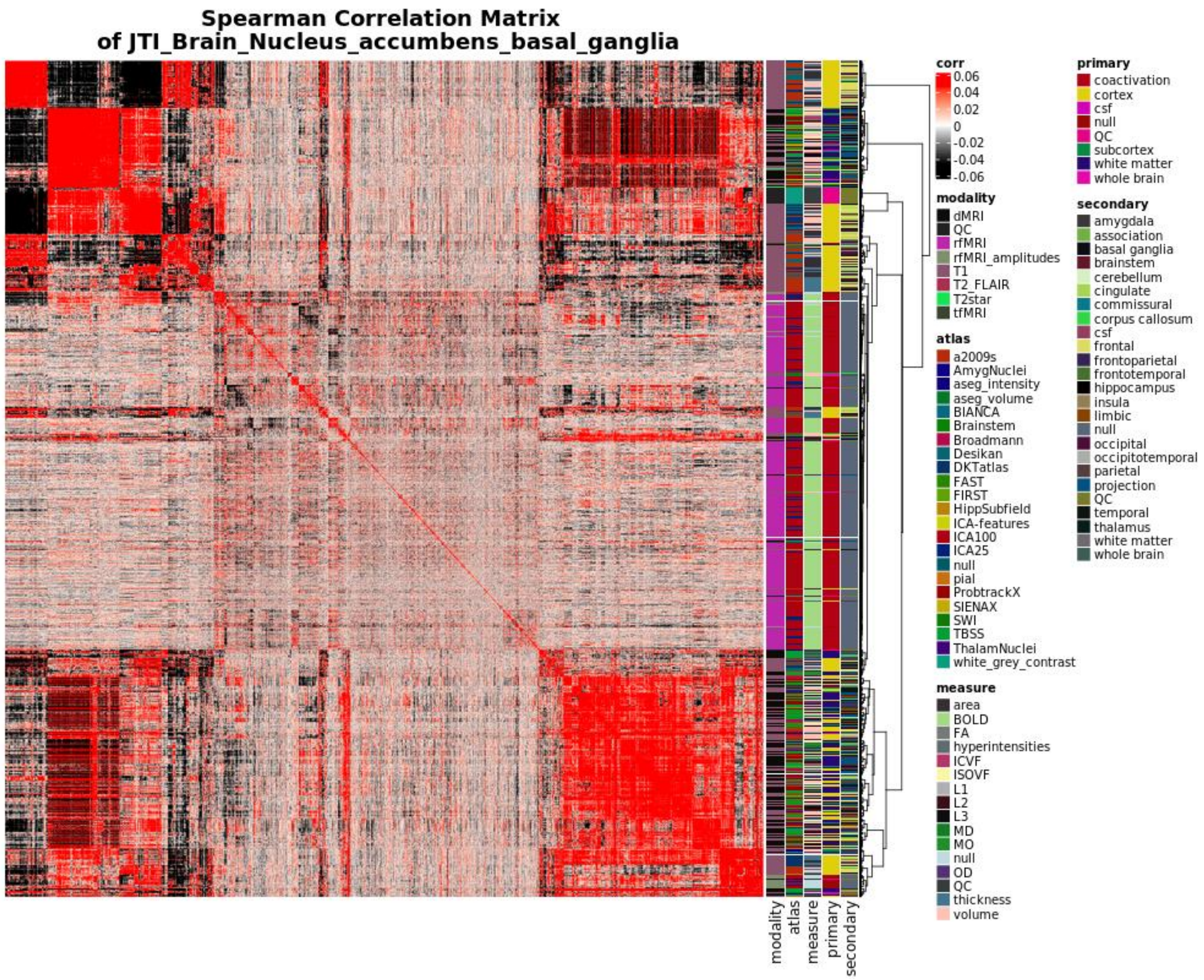


**Supplementary Figure 18:** We demonstrate NIPD similarity on the basis of GReX association effect size with a matrix detailing the pairwise spearman correlation distance between all NIDPs according to the unfiltered z-scores of all measured GReX associations as predicted by the nucleus accumbens. Annotations describe the MR imaging modality (modality), the atlas used in the extraction of pre-defined neurologic regions (atlas), the type of measurement characterized by each NIDP (measure), the type of brain tissue/MR element being examined (primary), and the named region of the brain characterized by the NIDP (secondary). These values are derived from the UKB supplementary NIDP information and detailed in Supplementary Table 1. Corresponding ranked ordering of NIDPs according to this tissue model can be found in Supplementary Table 12.

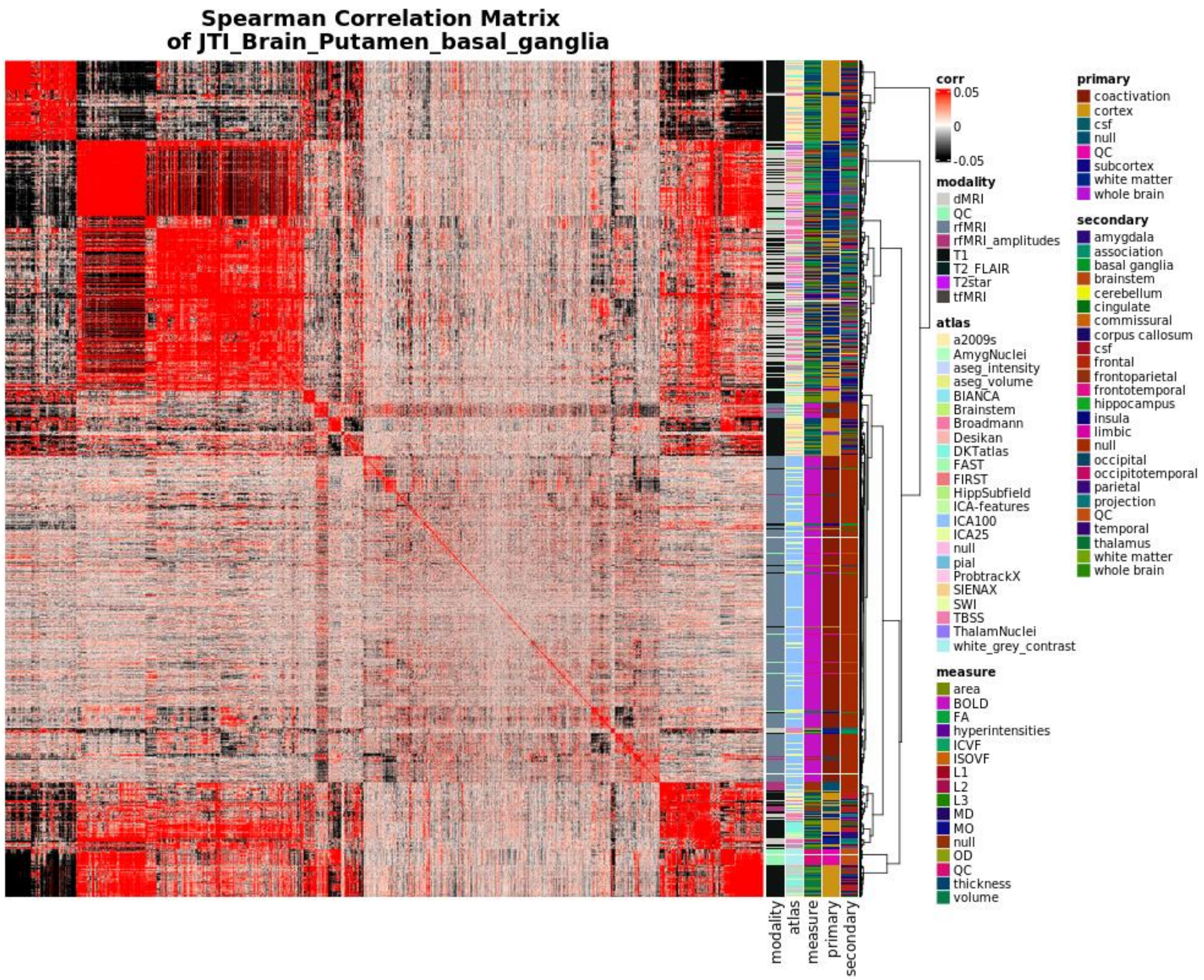


**Supplementary Figure 19:** We demonstrate NIPD similarity on the basis of GReX association effect size with a matrix detailing the pairwise spearman correlation distance between all NIDPs according to the unfiltered z-scores of all measured GReX associations as predicted by the putamen. Annotations describe the MR imaging modality (modality), the atlas used in the extraction of pre-defined neurologic regions (atlas), the type of measurement characterized by each NIDP (measure), the type of brain tissue/MR element being examined (primary), and the named region of the brain characterized by the NIDP (secondary). These values are derived from the UKB supplementary NIDP information and detailed in Supplementary Table 1. Corresponding ranked ordering of NIDPs according to this tissue model can be found in Supplementary Table 13.

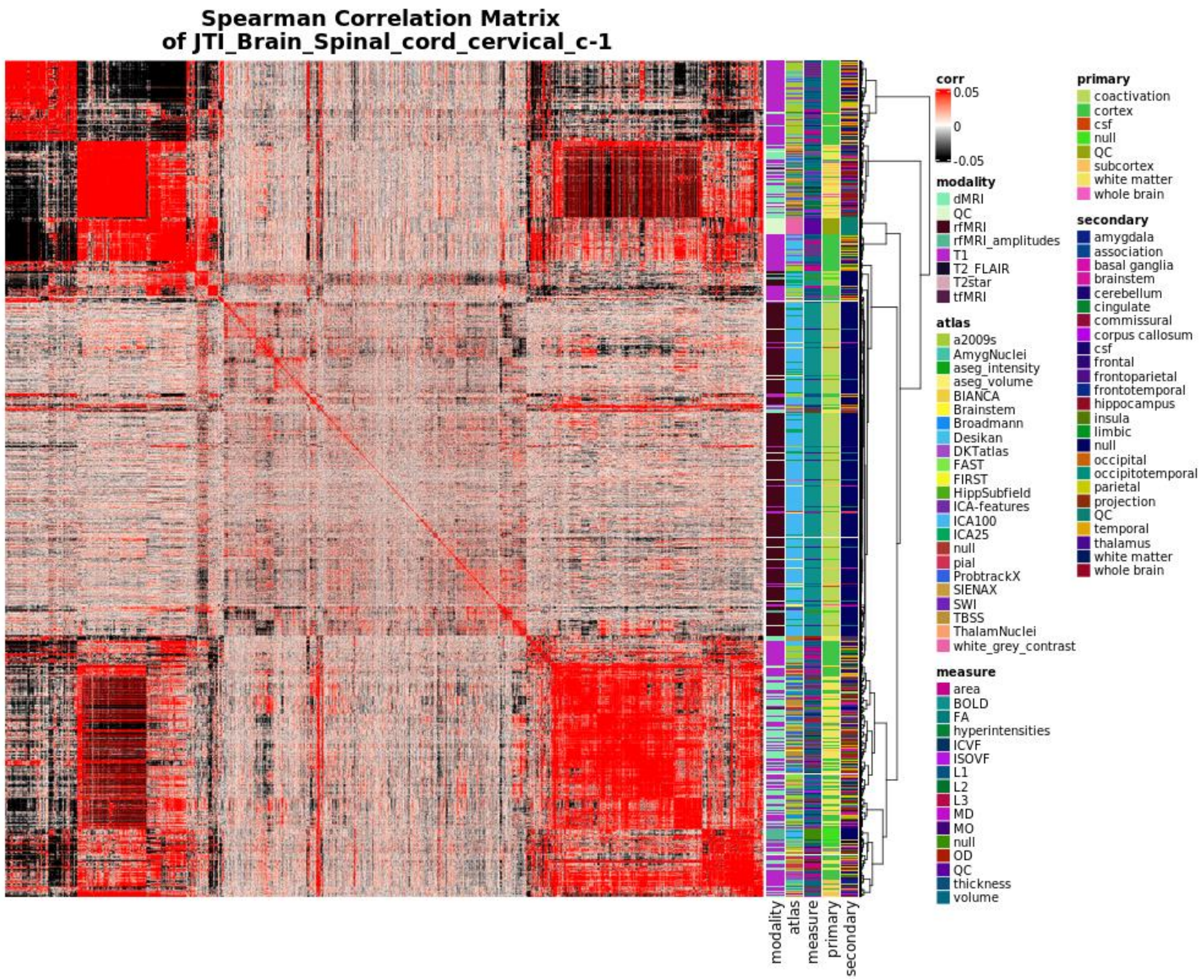


**Supplementary Figure 20:** We demonstrate NIPD similarity on the basis of GReX association effect size with a matrix detailing the pairwise spearman correlation distance between all NIDPs according to the unfiltered z-scores of all measured GReX associations as predicted by the spinal cord. Annotations describe the MR imaging modality (modality), the atlas used in the extraction of pre-defined neurologic regions (atlas), the type of measurement characterized by each NIDP (measure), the type of brain tissue/MR element being examined (primary), and the named region of the brain characterized by the NIDP (secondary). These values are derived from the UKB supplementary NIDP information and detailed in Supplementary Table 1. Corresponding ranked ordering of NIDPs according to this tissue model can be found in Supplementary Table 14.

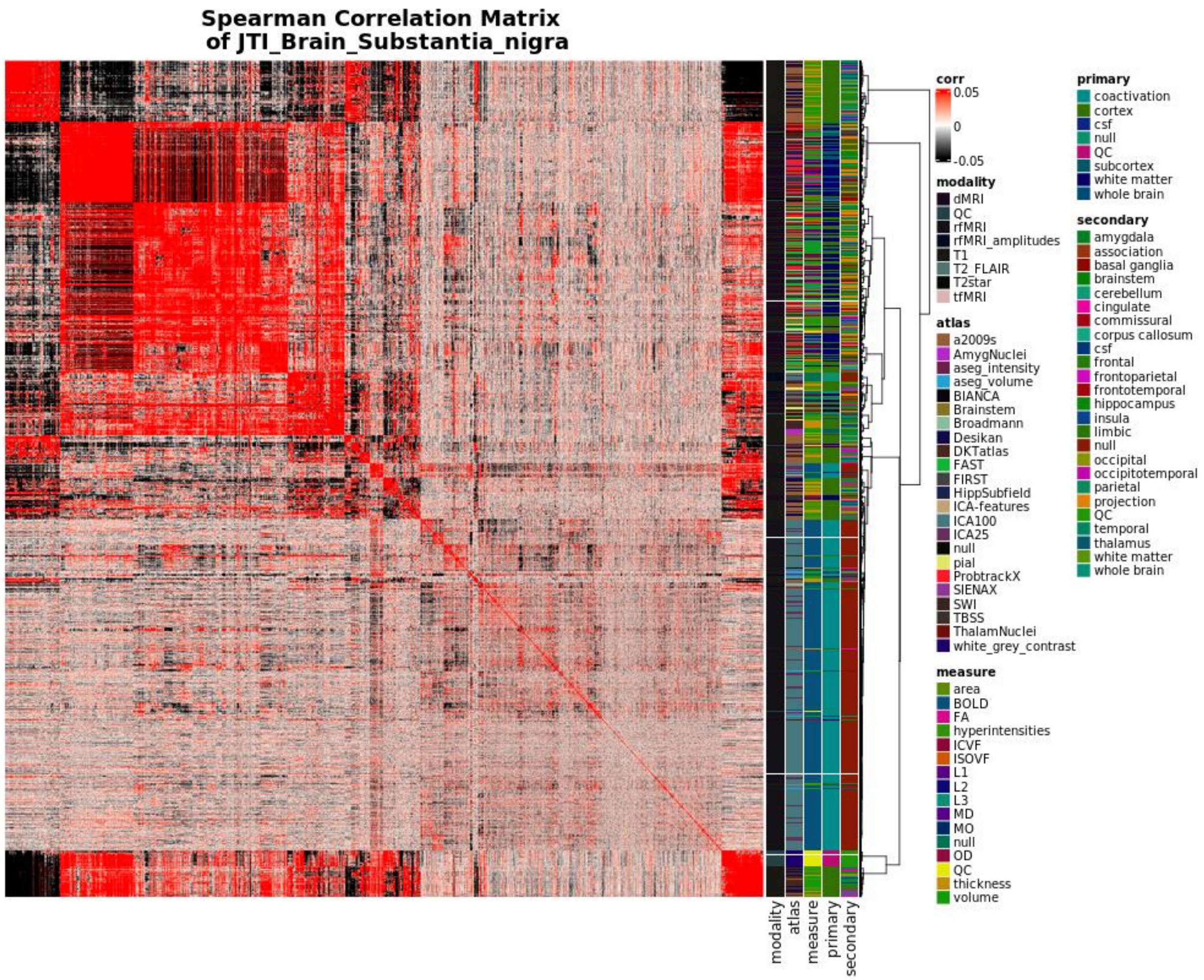


**Supplementary Figure 21:** We demonstrate NIPD similarity on the basis of GReX association effect size with a matrix detailing the pairwise spearman correlation distance between all NIDPs according to the unfiltered z-scores of all measured GReX associations as predicted by the substantia nigra. Annotations describe the MR imaging modality (modality), the atlas used in the extraction of pre-defined neurologic regions (atlas), the type of measurement characterized by each NIDP (measure), the type of brain tissue/MR element being examined (primary), and the named region of the brain characterized by the NIDP (secondary). These values are derived from the UKB supplementary NIDP information and detailed in Supplementary Table 1. Corresponding ranked ordering of NIDPs according to this tissue model can be found in Supplementary Table 15.

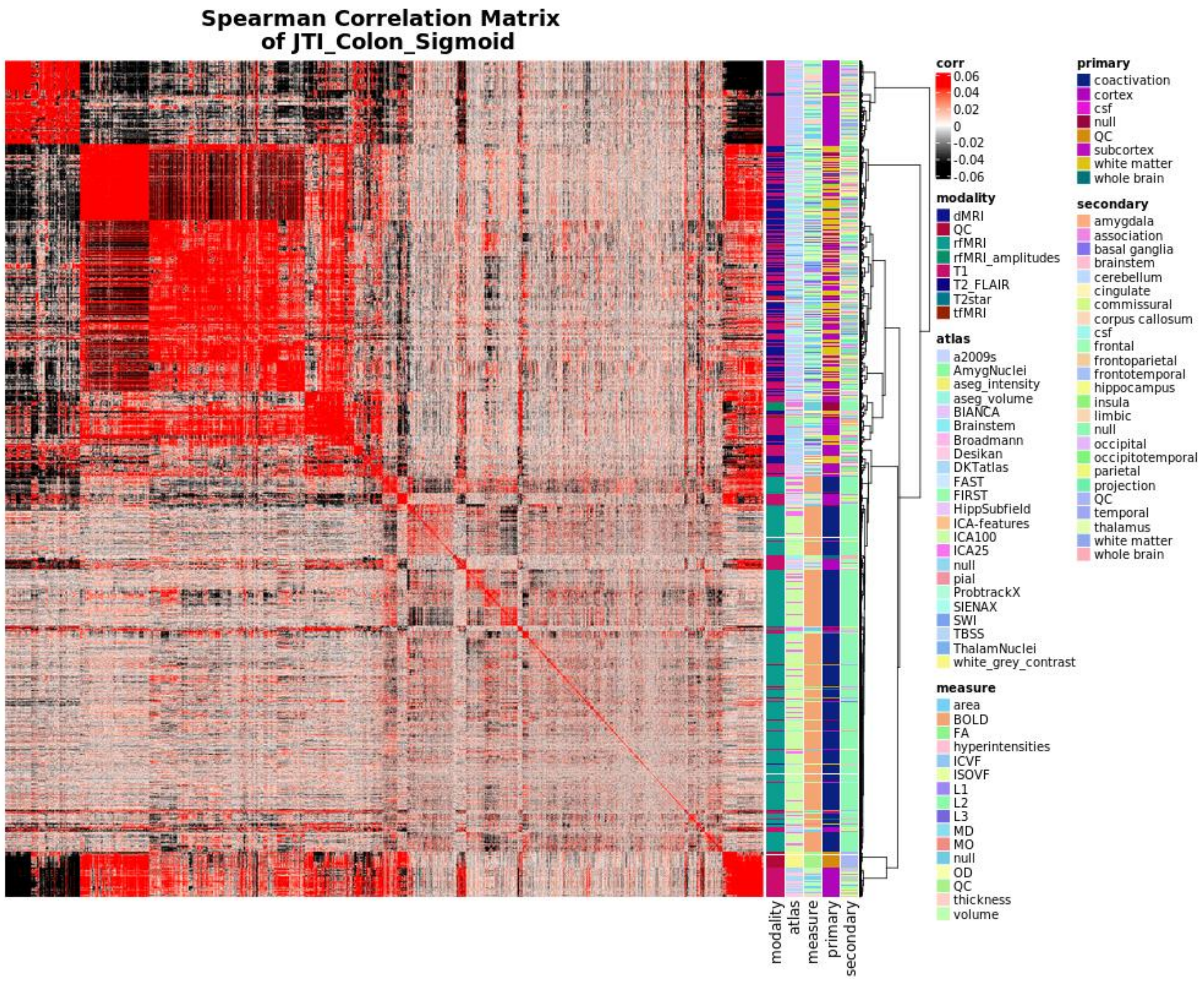


**Supplementary Figure 22:** We demonstrate NIPD similarity on the basis of GReX association effect size with a matrix detailing the pairwise spearman correlation distance between all NIDPs according to the unfiltered z-scores of all measured GReX associations as predicted by the sigmoid colon. Annotations describe the MR imaging modality (modality), the atlas used in the extraction of pre-defined neurologic regions (atlas), the type of measurement characterized by each NIDP (measure), the type of brain tissue/MR element being examined (primary), and the named region of the brain characterized by the NIDP (secondary). These values are derived from the UKB supplementary NIDP information and detailed in Supplementary Table 1. Corresponding ranked ordering of NIDPs according to this tissue model can be found in Supplementary Table 16.

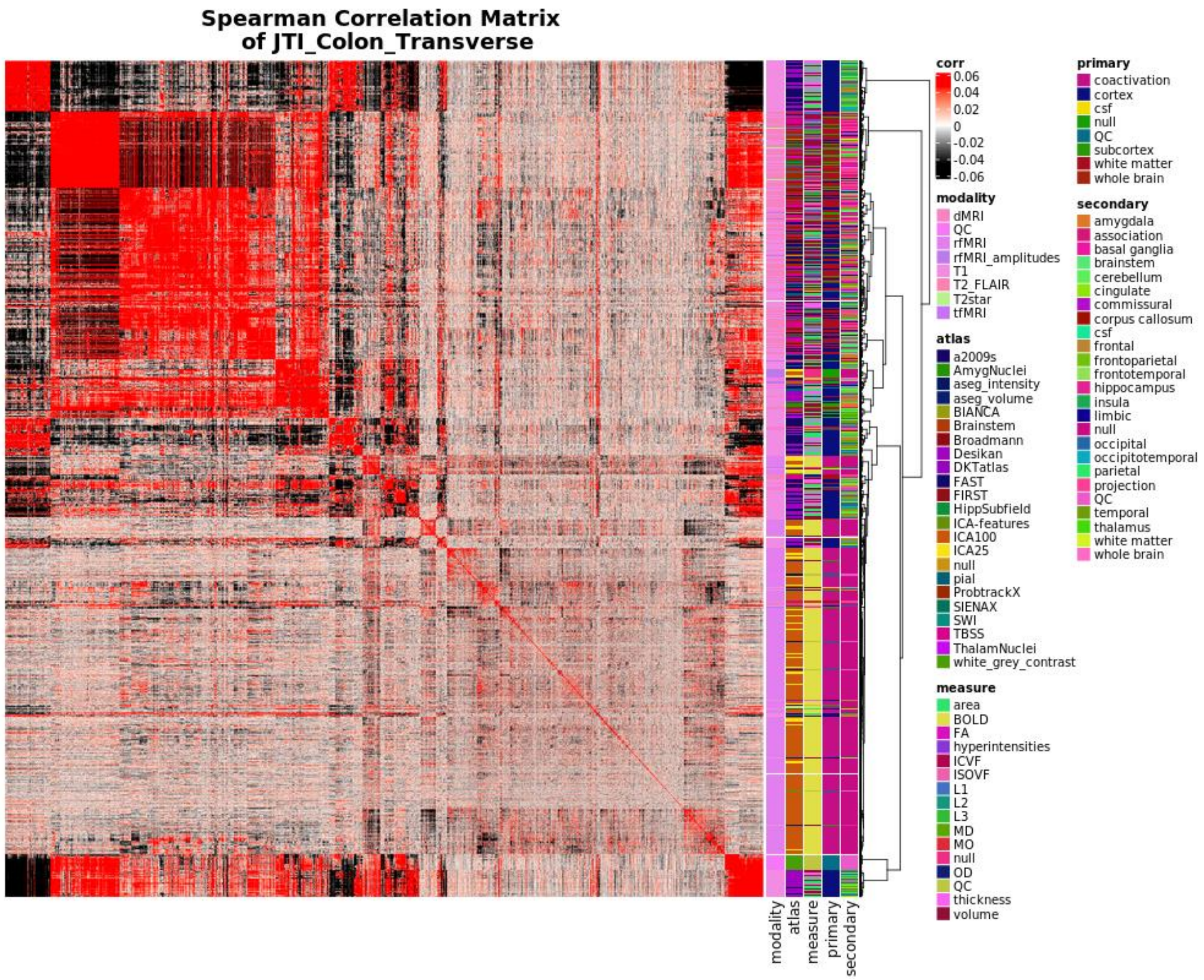


**Supplementary Figure 23:** We demonstrate NIPD similarity on the basis of GReX association effect size with a matrix detailing the pairwise spearman correlation distance between all NIDPs according to the unfiltered z-scores of all measured GReX associations as predicted by the transverse colon. Annotations describe the MR imaging modality (modality), the atlas used in the extraction of pre-defined neurologic regions (atlas), the type of measurement characterized by each NIDP (measure), the type of brain tissue/MR element being examined (primary), and the named region of the brain characterized by the NIDP (secondary). These values are derived from the UKB supplementary NIDP information and detailed in Supplementary Table 1. Corresponding ranked ordering of NIDPs according to this tissue model can be found in Supplementary Table 17.

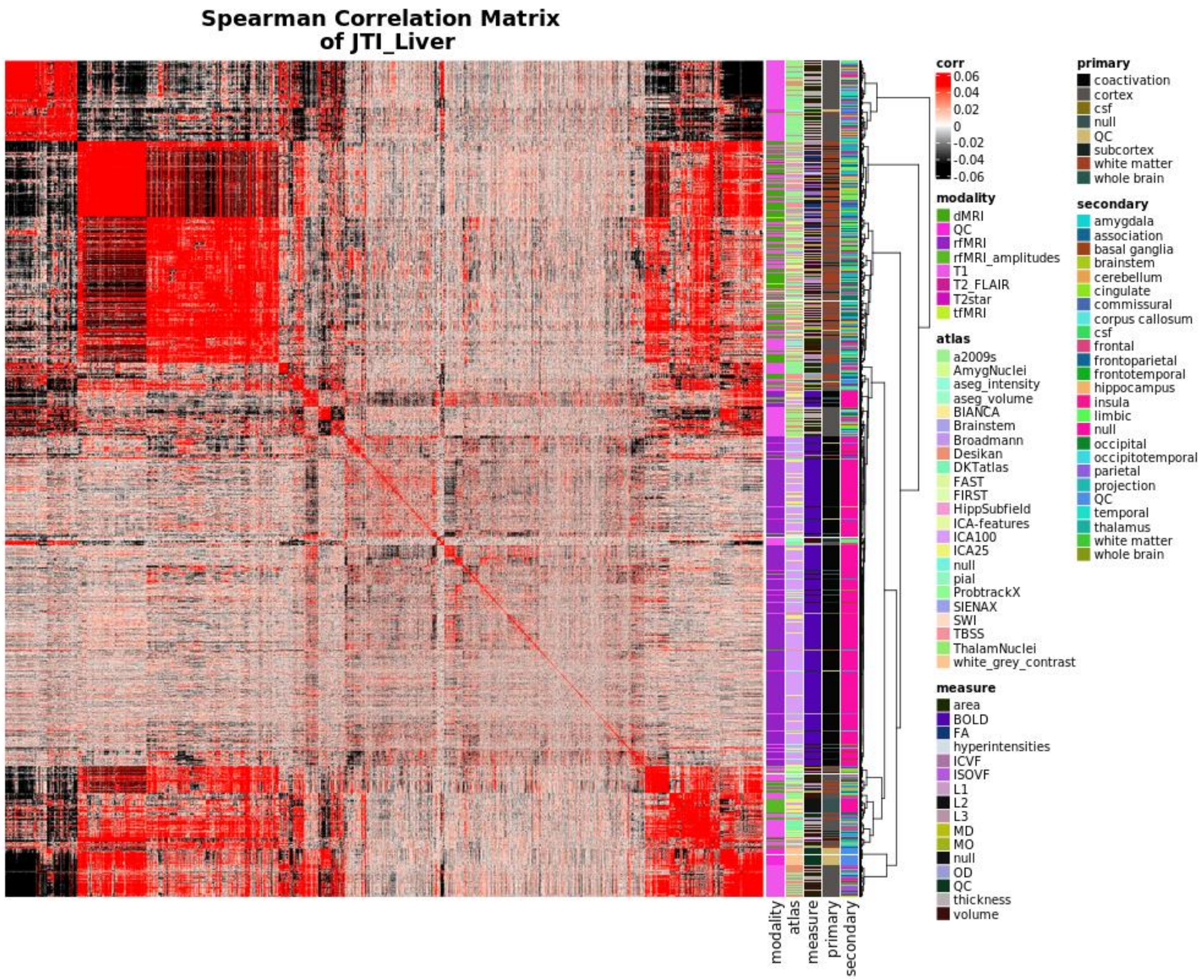


**Supplementary Figure 24:** We demonstrate NIPD similarity on the basis of GReX association effect size with a matrix detailing the pairwise spearman correlation distance between all NIDPs according to the unfiltered z-scores of all measured GReX associations as predicted by the liver. Annotations describe the MR imaging modality (modality), the atlas used in the extraction of pre-defined neurologic regions (atlas), the type of measurement characterized by each NIDP (measure), the type of brain tissue/MR element being examined (primary), and the named region of the brain characterized by the NIDP (secondary). These values are derived from the UKB supplementary NIDP information and detailed in Supplementary Table 1. Corresponding ranked ordering of NIDPs according to this tissue model can be found in Supplementary Table 18.

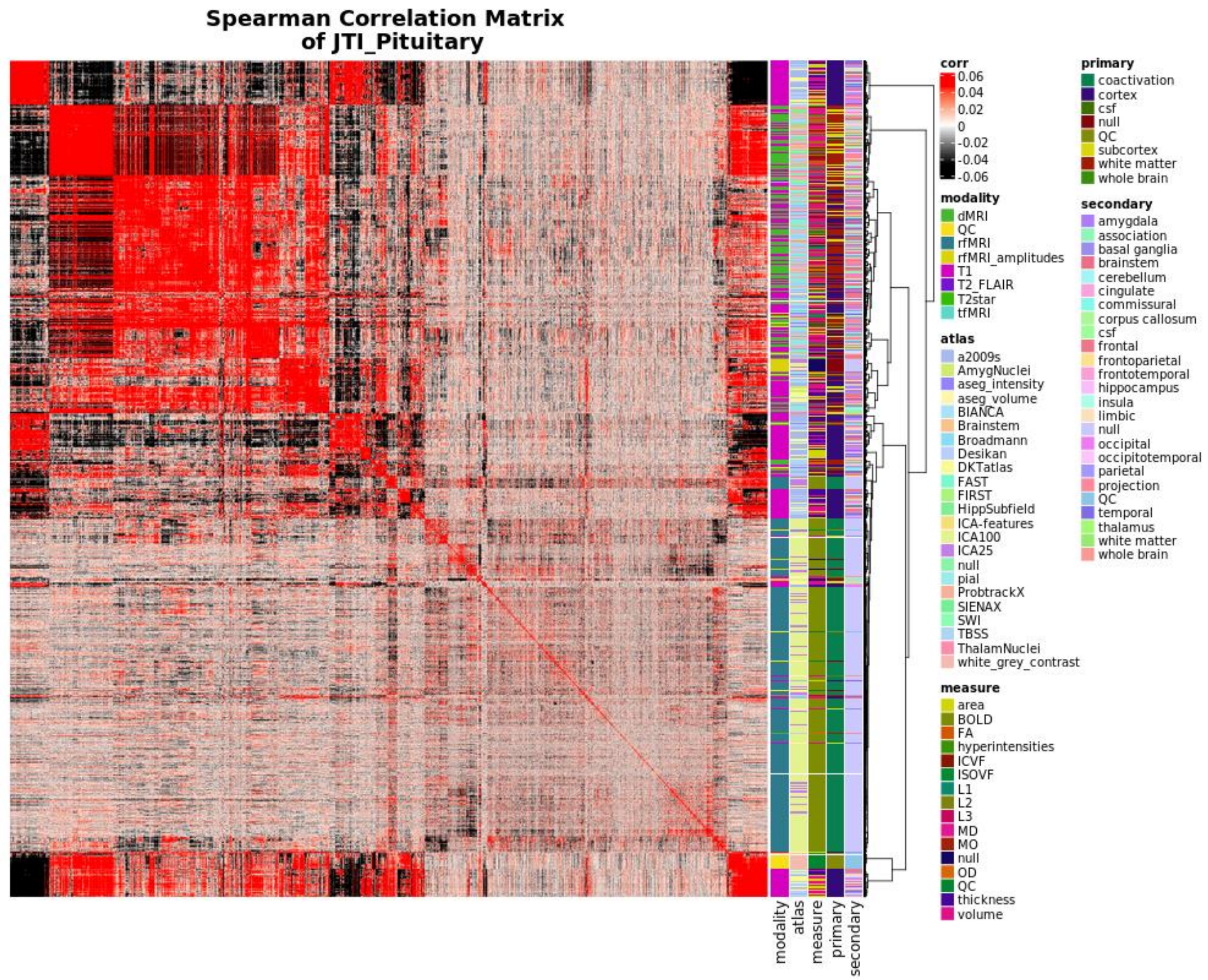


**Supplementary Figure 25:** We demonstrate NIPD similarity on the basis of GReX association effect size with a matrix detailing the pairwise spearman correlation distance between all NIDPs according to the unfiltered z-scores of all measured GReX associations as predicted by the pituitary. Annotations describe the MR imaging modality (modality), the atlas used in the extraction of pre-defined neurologic regions (atlas), the type of measurement characterized by each NIDP (measure), the type of brain tissue/MR element being examined (primary), and the named region of the brain characterized by the NIDP (secondary). These values are derived from the UKB supplementary NIDP information and detailed in Supplementary Table 1. Corresponding ranked ordering of NIDPs according to this tissue model can be found in Supplementary Table 19.

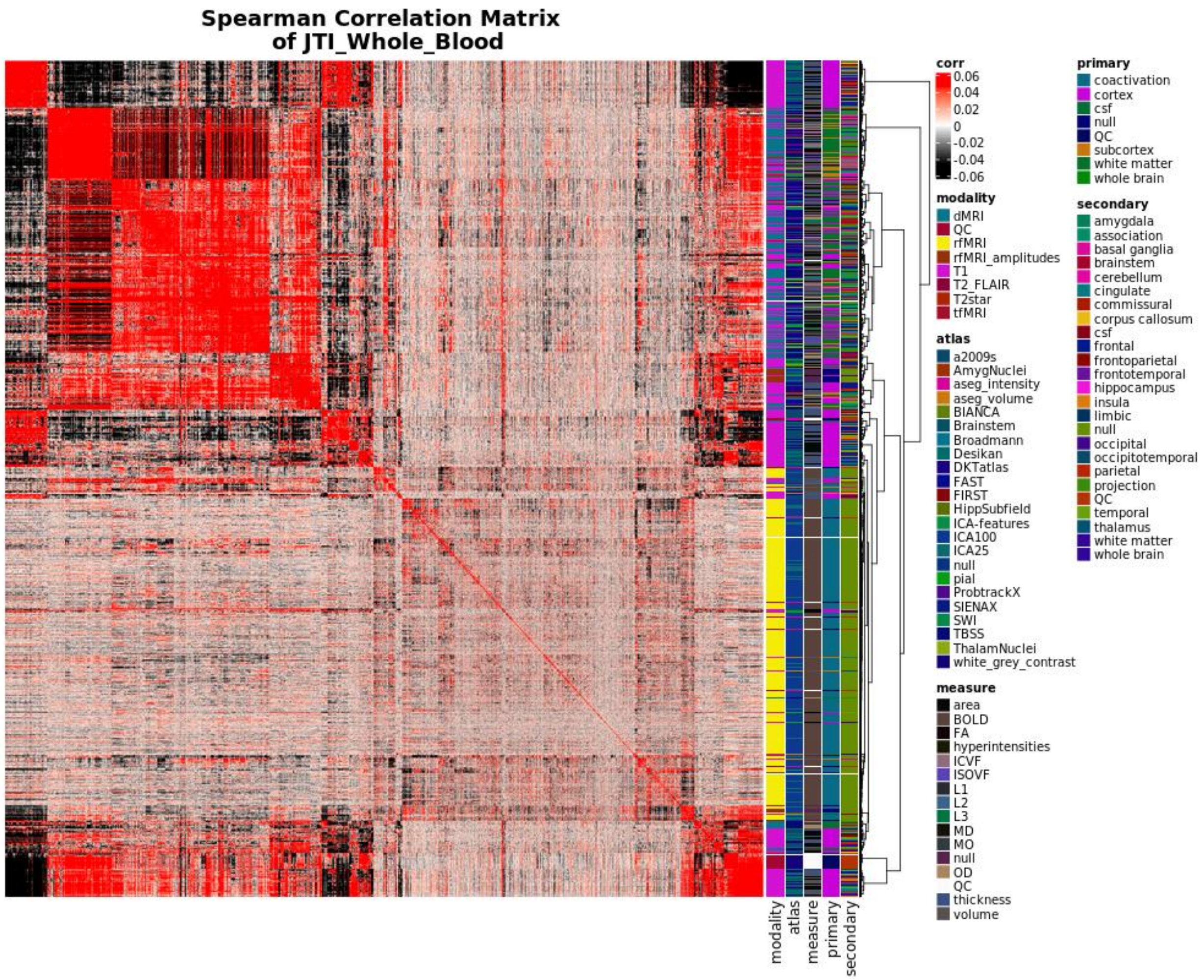


**Supplementary Figure 26:** We demonstrate NIPD similarity on the basis of GReX association effect size with a matrix detailing the pairwise spearman correlation distance between all NIDPs according to the unfiltered z-scores of all measured GReX associations as predicted by the whole blood. Annotations describe the MR imaging modality (modality), the atlas used in the extraction of pre-defined neurologic regions (atlas), the type of measurement characterized by each NIDP (measure), the type of brain tissue/MR element being examined (primary), and the named region of the brain characterized by the NIDP (secondary). These values are derived from the UKB supplementary NIDP information and detailed in Supplementary Table 1. Corresponding ranked ordering of NIDPs according to this tissue model can be found in Supplementary Table 20.

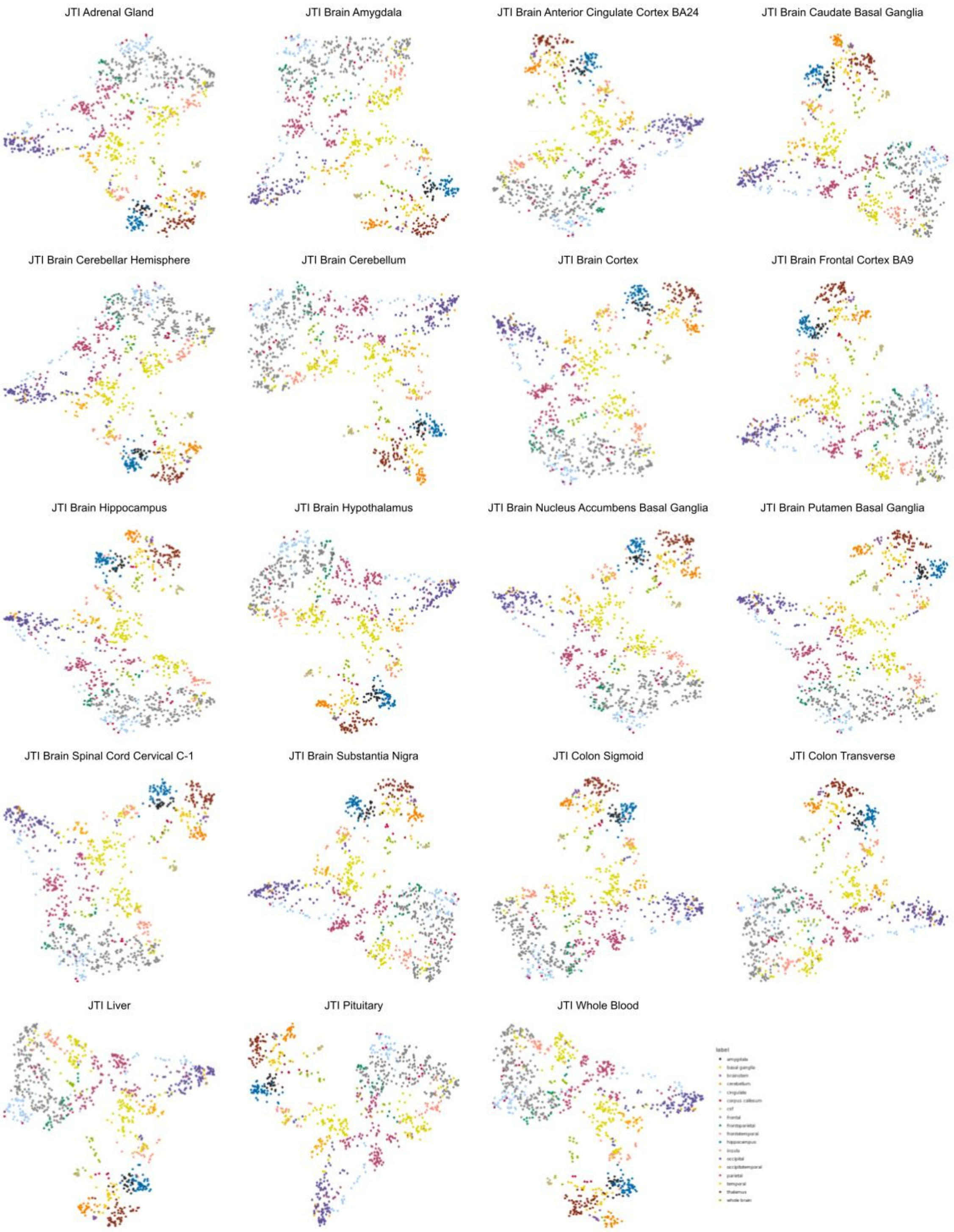


**Supplementary Figure 27:** UMAP projection of T1 derived NIDPs corresponding to cortical surface area and volumes according to all 19 JTI tissue models. The distribution of points in the cartesian plane is derived exclusively from Euclidean distance between the predicted GReX effect size vectors for each NIDP. NIDPs are colored according to cortical region.

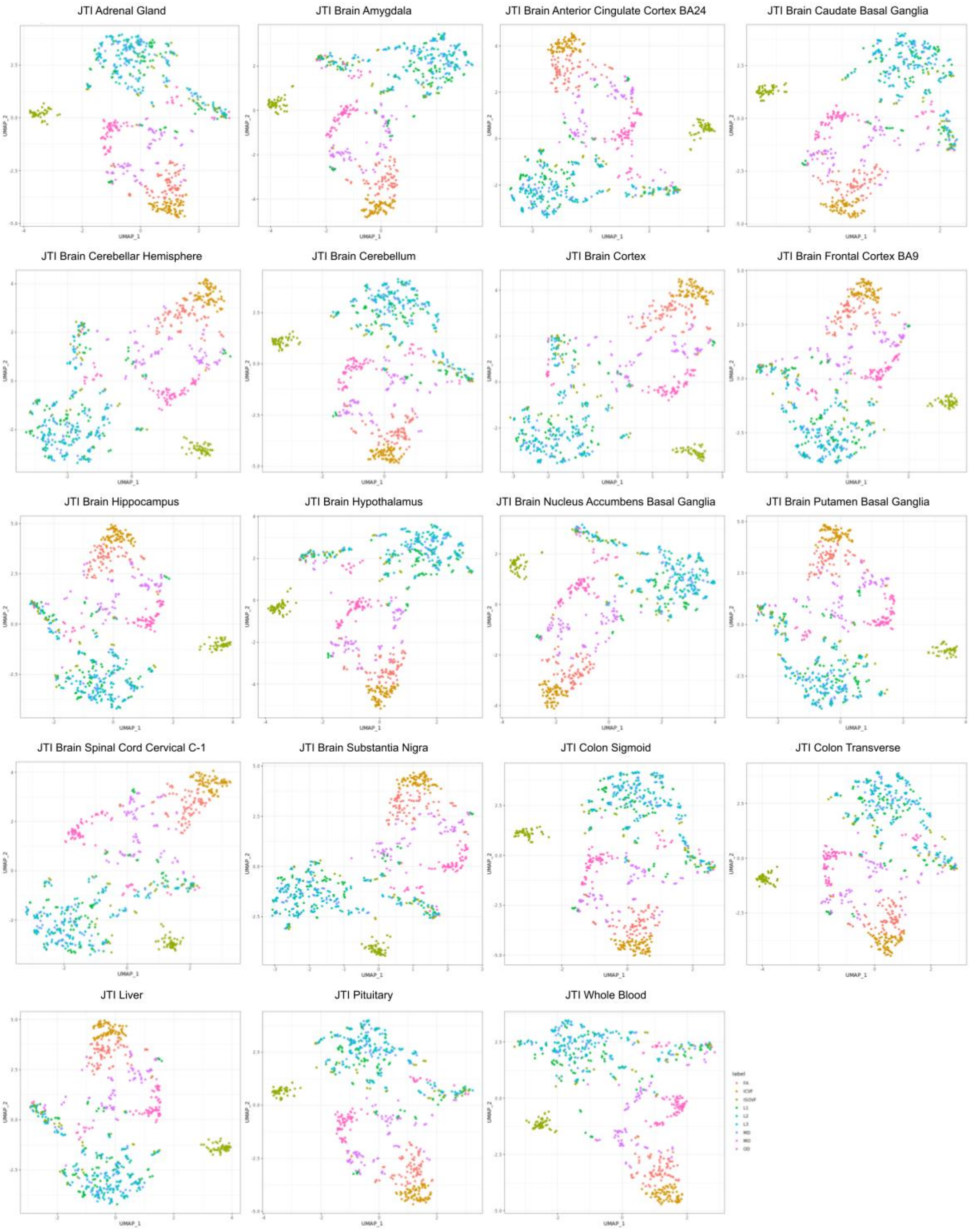


**Supplementary Figure 28:** UMAP projection of all dMRI-derived NIDPs according to all 19 JTI tissue models. The distribution of points in the cartesian plane is derived exclusively from Euclidean distance between the predicted GReX effect size vectors for each NIDP. NIDPs are colored according to tractography measures. **FA:** Fractional Anisotropy, **ICVF:** Intra-cellular Volume Fraction, **ISOVF:** Isovolumetric Volume Fraction, **L1-3:** Principle diffusion direction (global), **MD:** Mean Diffusivity, **MO:** Mode of Anisotropy, **OD:** Orbital Diffusivity

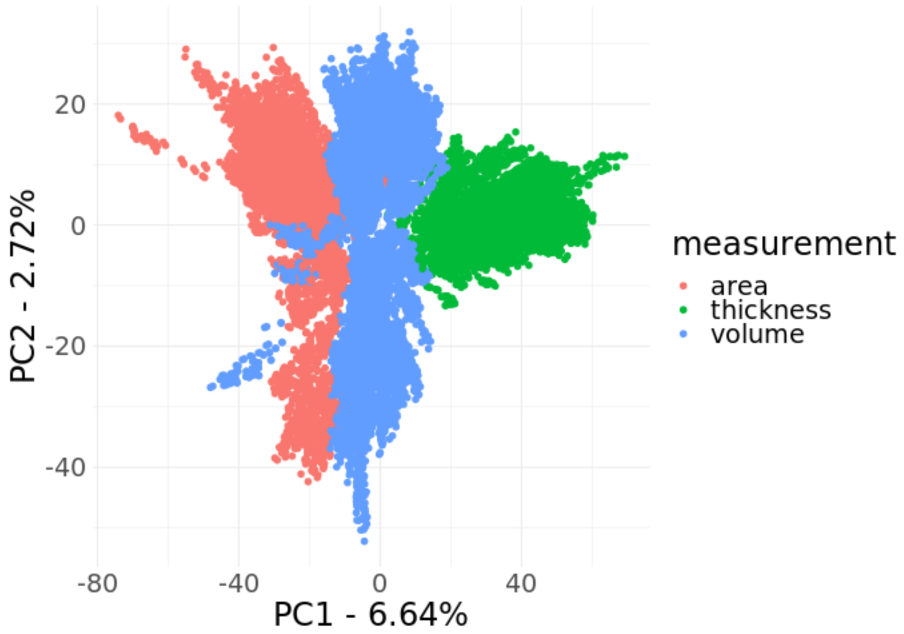

**Supplementary Figure 29:** PCA projection of all T1 derived NIDPs corresponding to cortical surface area, thickness, and volumes according to all 19 JTI tissue models. NIDPs are colored according to cortical measure.

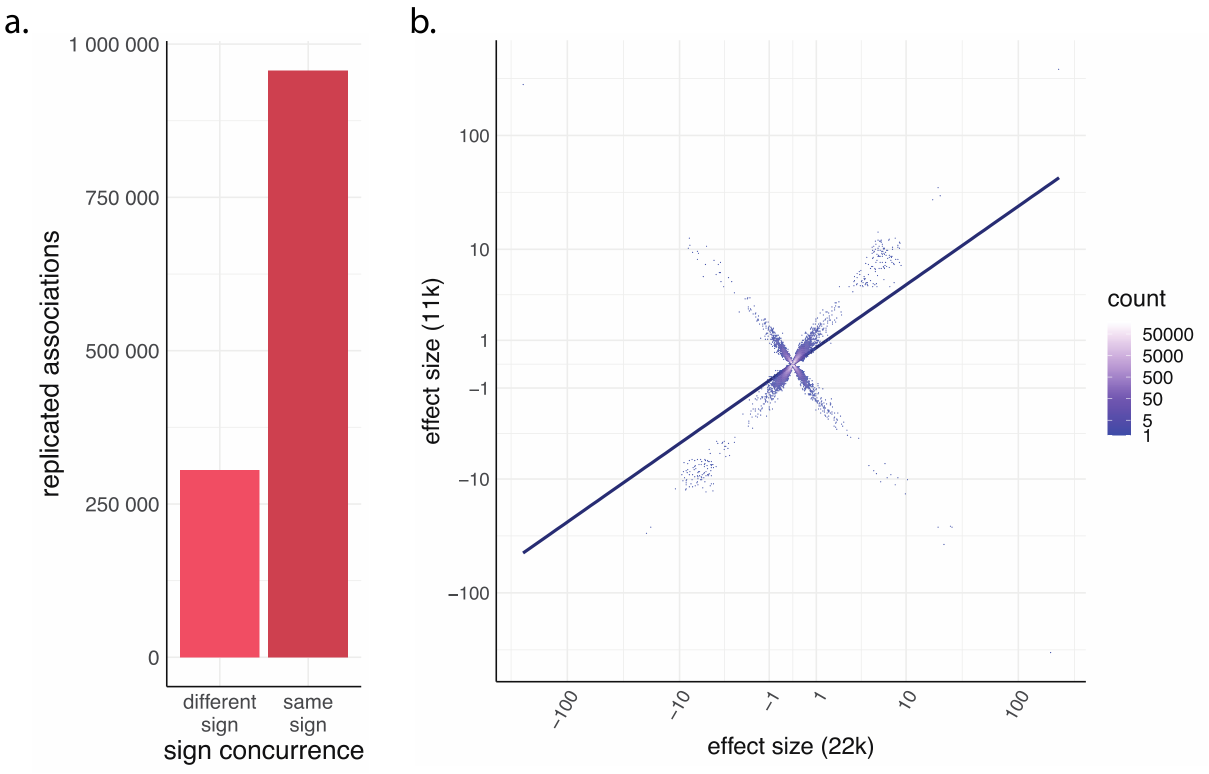


**Supplementary Figure 30:** Internal replication of neuroimaging TWAS associations by effect size using a nominal p-value significance threshold of 0.05.
a. Relative counts of GReX-NIDP associations present in neuroimaging TWAS data from 11K subjects and neuroimaging TWAS data from 22K subjects, which share the same direction of effect vs those that show differing directions of effect.

b. The correlation between effect sizes of tissue-concordant GReX-NIDP associations shared across the neuroimaging TWAS data from 11K subjects and the neuroimaging TWAS data from 22K subjects. Data are plotted according to a pseudolog10 function with point color representing degree of overlap at each graphical coordinate. The trend line reflects linear regression. Spearman correlation = 0.547, p-value < 2.2e-16.

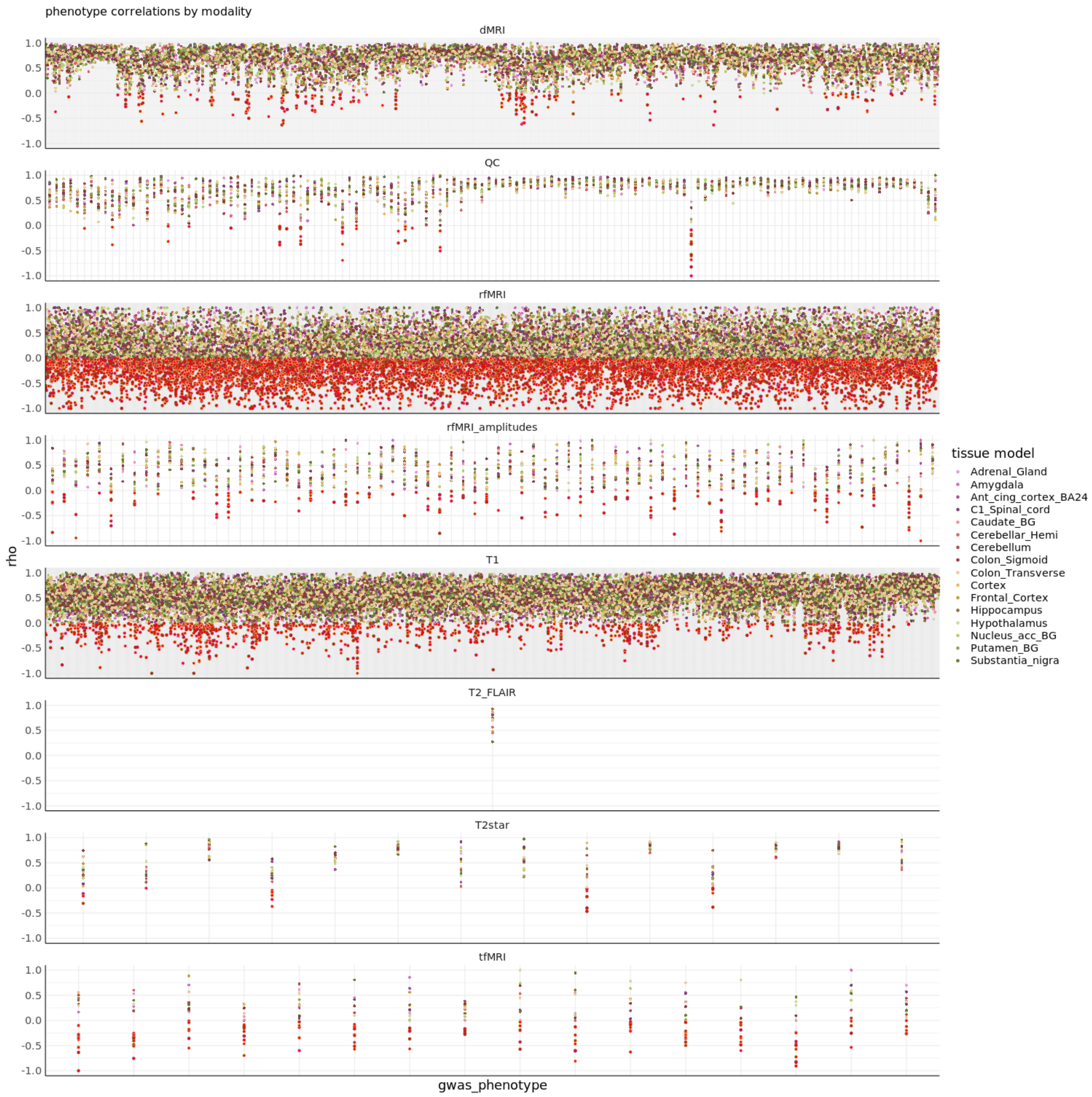


**Supplementary Figure 31:** We show the correlation between effect sizes of tissue-concordant GReX-NIDP associations shared across the neuroimaging TWAS data from 11K subjects and the neuroimaging TWAS data from 22K subjects. All data is filtered by a nominal p-value TWAS threshold of 0.05. The data specifically show the distribution of all NIDPs plotted according to the spearman correlation coefficient between gene sets as calculated from TWAS in the 11k and 22K data subsets. Panels represent imaging modalities. Point color represents the tissue model with all points showing anti-correlation filled red. (rho < 0)

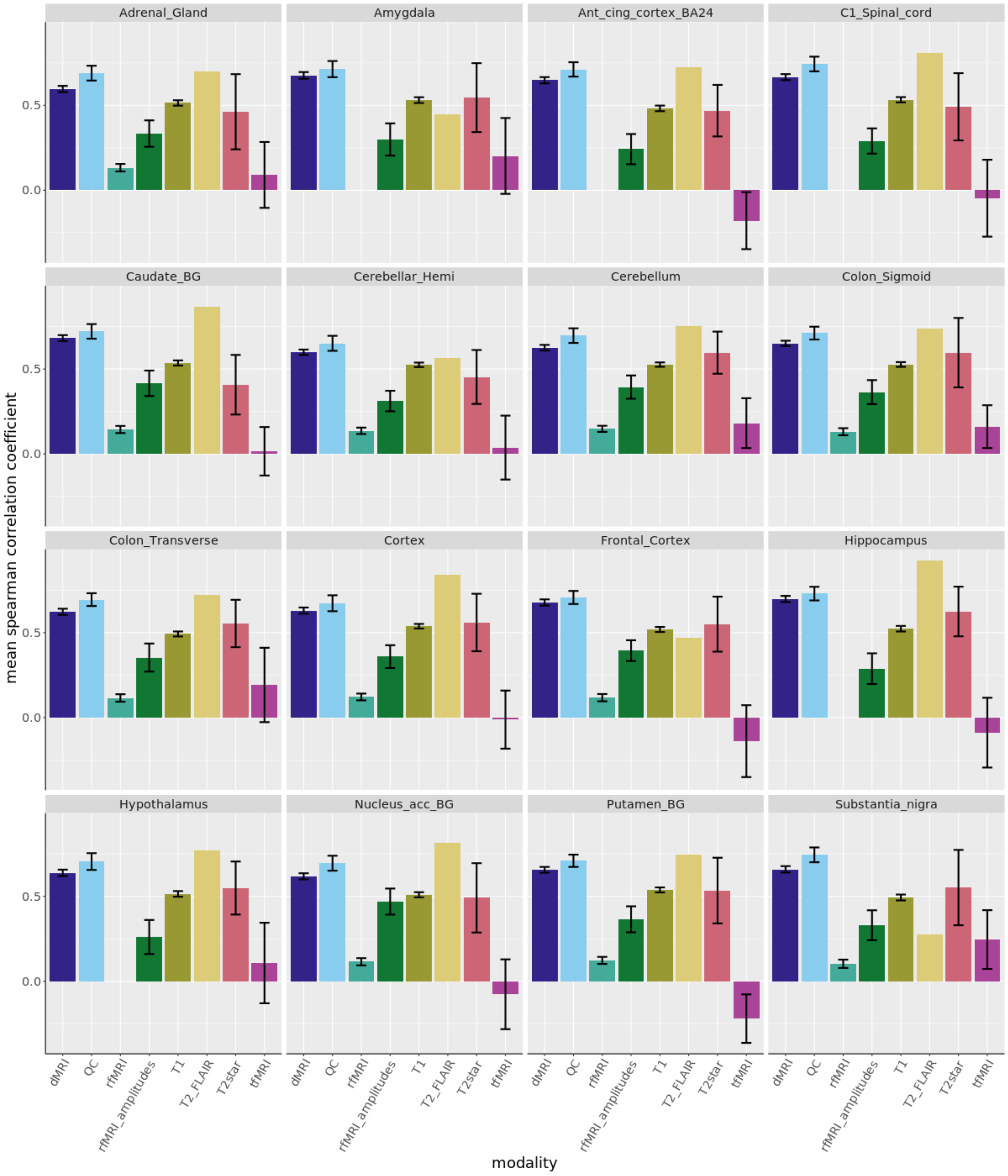


**Supplementary Figure 32:** Quantification of data replication across 11K and 22K data subsets in the Neuroimaging TWAS. For all the NIDPs with GReX associations that met a nominal p-value threshold in both the 11K and 22K subsets of data I calculated the mean spearman correlation. I then grouped these correlations according to the tissue model and modality and took the mean correlation across all NIDPs. These are plotted above with error bars representing the 95% confidence interval.

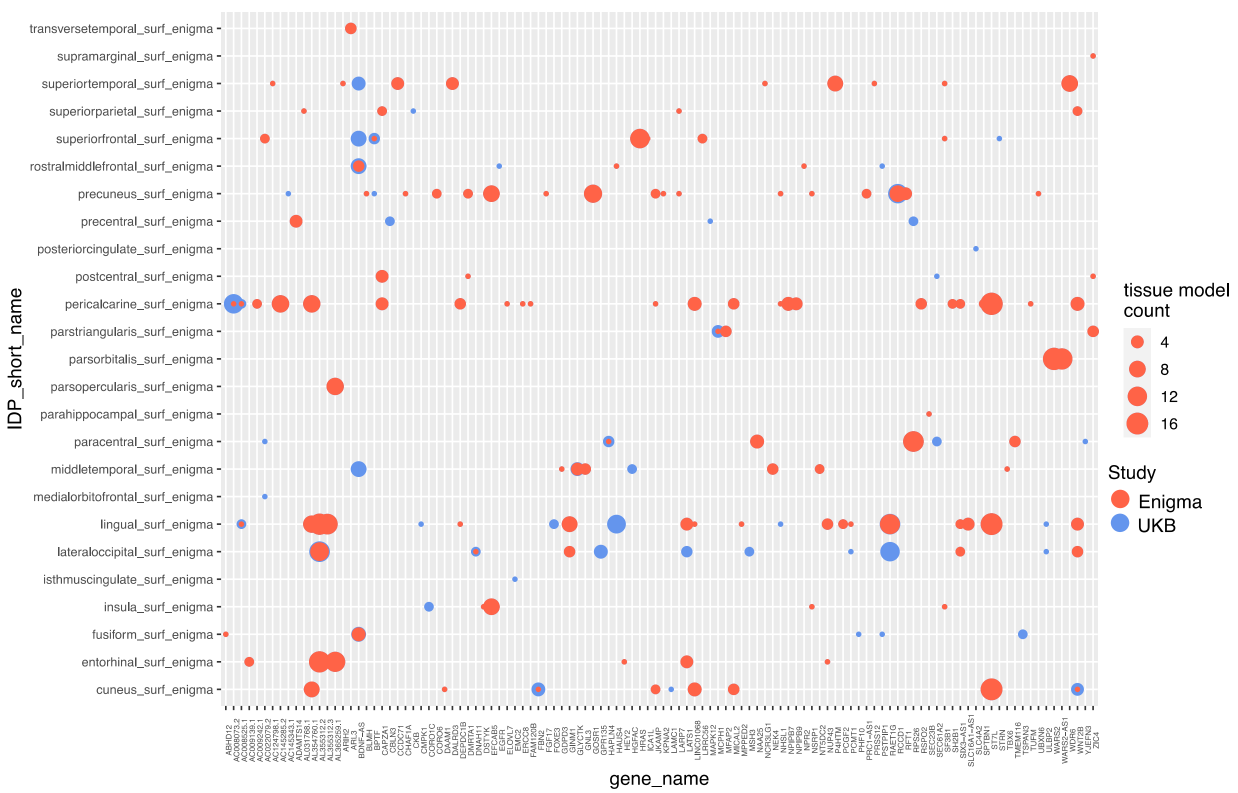


**Supplementary Figure 33:** Replication analysis of NeuroimaGene TWAS of cortical Surface Area data compared to TWAS data derived from an external cortical GWAS conducted by the ENIGMA consortium. The vertical axis describes NIDPs while the horizontal atlas details gene expression. The size of the points represents the number of tissue models in which the analysis reached FDR significance in the discovery cohort ( blue) and the size of the red points represents the number of replicated associations according to a nominal threshold in the ENIGMA data. Points which appear fully red represent full replication of all UKB associations in all tissue models in the ENIGMA replication cohort.

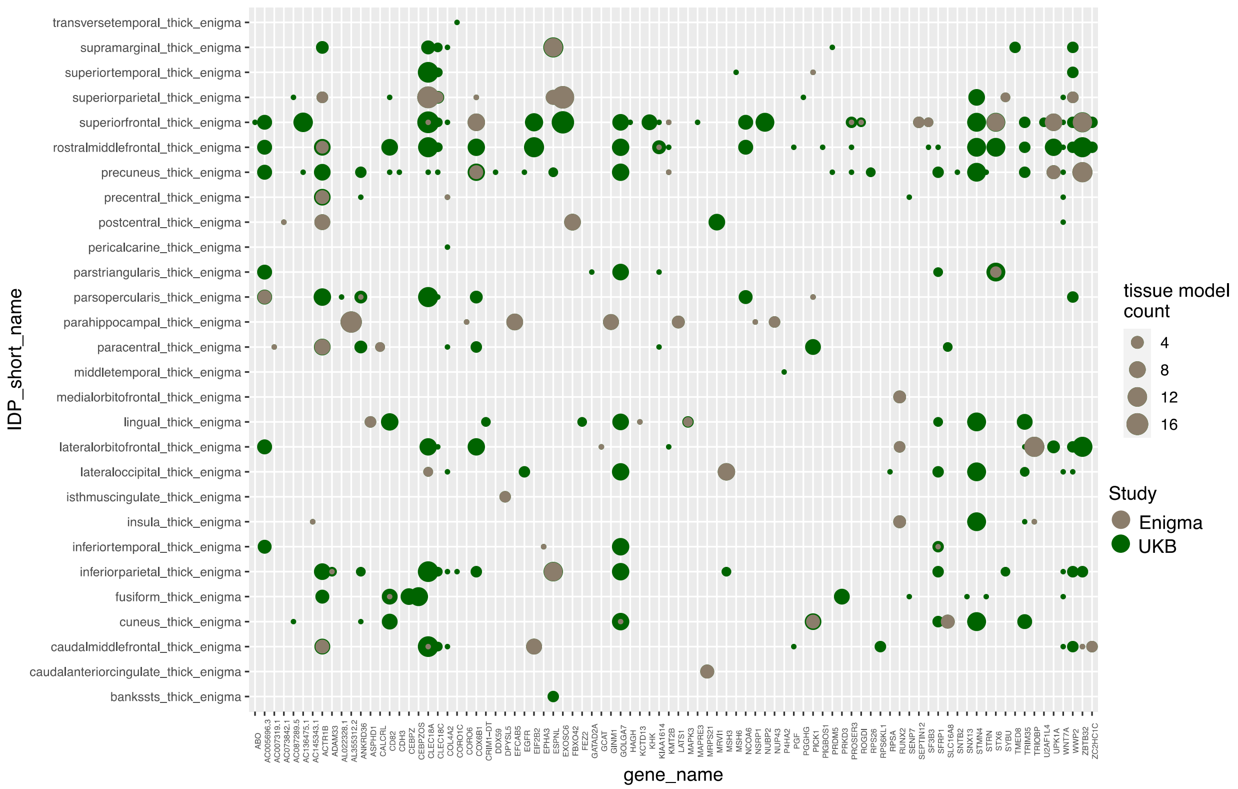


**Supplementary Figure 34:** Replication analysis of NeuroimaGene TWAS of cortical thickness data compared to TWAS data derived from an external cortical GWAS conducted by the ENIGMA consortium. The vertical axis describes NIDPs while the horizontal atlas details gene expression. The size of the points represents the number of tissue models in which the analysis reached FDR significance in the discovery cohort (green) and the size of the grey points represents the number of replicated associations according to a nominal threshold in the ENIGMA data. Points which appear fully red represent full replication of all UKB associations in all tissue models in the ENIGMA replication cohort.

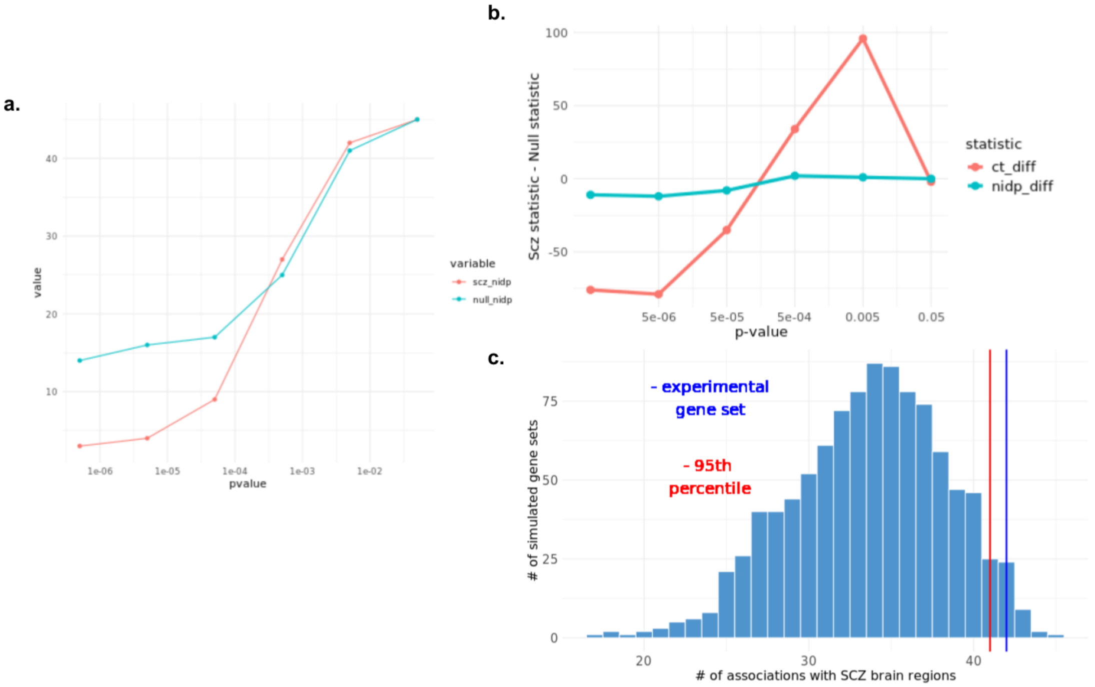


**Supplementary Figure 35:** NIDP specific p-value optimization from SCZ analysis. **a.** Comparison of unique NIDPs in the set of associations between SCZ GReX measures and SCZ NIDPs (scz_nidp) and unique NIDPs between random GReX measures and SCZ NIDPs (null_nidps) at different p-value thresholds. **b.** Difference plot detailing the total number of associations and unique nidps in those associations between SCZ GReX measures and SCZ NIDPs (scz_ct/ scz_nidp) minus the total number of associations between random GReX measures and SCZ NIDPs (null_ct/ null_nidp) at different p-value thresholds. **c.** Quantification of unique NIDPs in SCZ GReX associations with SCZ-NIDPs compared to a null distribution at p-value <= 0.005. The experimental gene set represents the set of GReX measures associated with SCZ according to Trubetskoy et al. The histogram represents the distribution of unique NIDPs in associations between 1000 random equally sized gene sets and SCZ-NIDPs. The line in red represents the limit of NIDP counts exceeded by only 0.05 of the random gene sets. The blue line represents the number of unique NIDPs in associations between SCZ-associated endophenotypes.

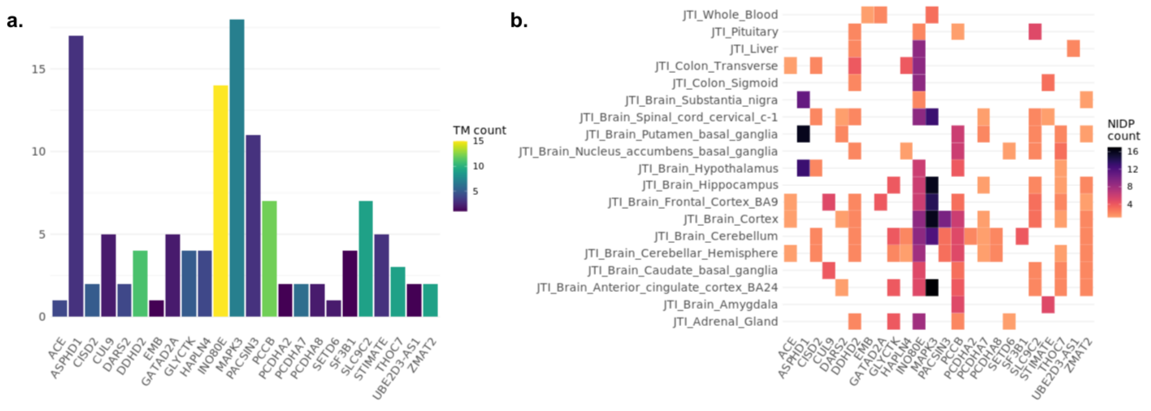


**Supplementary Figure 36:** Supporting figures for Schizophrenia image-directed analyses. **a.** Distribution of GReX-NIDP associations from the image directed schizophrenia analysis. The X-axis details GReX genes that are associated with SCZ in Trubetskoy et al. The Y-axis represents the number of cortical and subcortical NIDPs from the ENIGMA schizophrenia neuroimaging study that each GReX measure is associated with. The color of each bar represents the number of JTI-enriched tissue-specific eQTL models in which each GReX-NIDP association was statistically significant by the optimized p-value and causally implicated via Mendelian Randomization. **b.** Distribution of MR-JTI significant associations between SCZ GReX measures and SCZ-associated NIDPs stratified by tissue context as defined by 19 JTI-enriched tissue models.

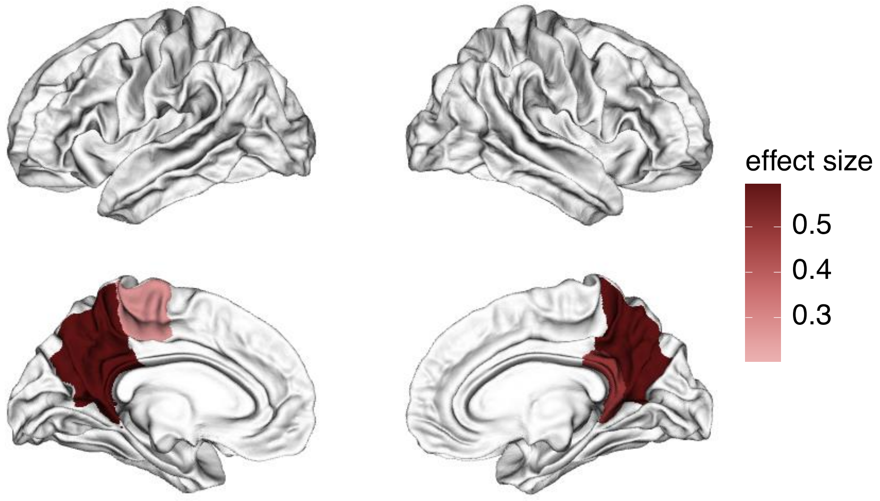


**Supplementary Figure 37:** Visualization of the cortical surface area measures implicated by GReX for schizophrenia-associated genes as detected in the Desikan Atlas using fsbrain. The effect size represents the mean GReX-NIDP effect size corrected for allelic heterogeneity via Mendelian randomization. The intensity of the effect size represents the magnitude of the effect size and the color represents the direction of effect of increased GReX on the size of the region in question (blue = positive, red = negative).
